# Determinants of pre-vaccination antibody responses to SARS-CoV-2: a population-based longitudinal study (COVIDENCE UK)

**DOI:** 10.1101/2021.11.02.21265767

**Authors:** Mohammad Talaei, Sian Faustini, Hayley Holt, David A. Jolliffe, Giulia Vivaldi, Matthew Greenig, Natalia Perdek, Sheena Maltby, Carola M Bigogno, Jane Symons, Gwyneth A Davies, Ronan A Lyons, Christopher J Griffiths, Frank Kee, Aziz Sheikh, Alex G Richter, Seif O Shaheen, Adrian R Martineau

## Abstract

**Background:** Prospective population-based studies investigating multiple determinants of pre-vaccination antibody responses to SARS-CoV-2 are lacking.

**Methods:** We did a prospective population-based study in SARS-CoV-2 vaccine-naive UK adults recruited between May 1 and November 2, 2020, without a positive swab test result for SARS-CoV-2 prior to enrolment. Information on 88 potential sociodemographic, behavioural, nutritional, clinical and pharmacological risk factors was obtained through online questionnaires, and combined IgG/IgA/IgM responses to SARS-CoV-2 spike glycoprotein were determined in dried blood spots obtained between November 6, 2020 and April 18, 2021. We used logistic and linear regression to estimate adjusted odds ratios (aORs) and adjusted geometric mean ratios (aGMRs) for potential determinants of SARS-CoV-2 seropositivity (all participants) and antibody titres (seropositive participants only), respectively.

**Results:** 1696 (15.2%) of 11,130 participants were seropositive. Factors independently associated with increased risk of SARS-CoV-2 seropositivity included frontline health/care occupation (aOR 1.86, 95% CI 1.48–2.33), international travel (1.20, 1.07–1.35), number of visits to shops and other indoor public places (≥5 vs. 0/week: 1.29, 1.06-1.57, P-trend=0.01), body mass index (BMI) ≥25 *vs* <25 kg/m^2^ (1.24, 1.11–1.39), Asian/Asian British *vs* White ethnicity (1.65, 1.10–2.49), and alcohol consumption ≥15 *vs* 0 units/week (1.23, 1.04–1.46). Light physical exercise associated with decreased risk (0.80, 0.70–0.93, for ≥10 *vs* 0–4 h/week). Among seropositive participants, higher titres of anti-Spike antibodies associated with factors including BMI ≥30 *vs* <25 kg/m^2^ (aGMR 1.10, 1.02–1.19), Asian/Asian British *vs* White ethnicity (1.22, 1.04–1.44), frontline health/care occupation (1.24, 95% CI 1.11–1.39), international travel (1.11, 1.05–1.16), and number of visits to shops and other indoor public places (≥5 vs. 0/week: 1.12, 1.02-1.23, P-trend=0.01); these associations were not substantially attenuated by adjustment for COVID-19 disease severity.

**Conclusions:** Higher alcohol consumption and reduced light physical exercise represent new modifiable risk factors for SARS-CoV-2 infection. Recognised associations between Asian/Asian British ethnic origin and obesity and increased risk of SARS-CoV-2 seropositivity were independent of other sociodemographic, behavioural, nutritional, clinical and pharmacological factors investigated. Among seropositive participants, higher titres of anti-Spike antibodies in people of Asian ancestry and in obese people were not explained by greater COVID-19 disease severity in these groups.

**Funding:** Barts Charity, Health Data Research UK.

## Introduction

The COVID-19 pandemic has caused more than 220 million recorded infections and over 4.5 million recorded deaths,^1^ with these figures representing only a portion of the true burden.^2^ Large, population-based studies have identified various risk factors for SARS-CoV-2 infection, including non-White ethnicity and lower educational attainment.^3–5^ However, the vast majority of studies have been based on routine real-time reverse transcription PCR (RT-PCR) testing in healthcare settings or in the community; consequently, they are potentially open to collider bias, as the probability of being tested for infection can itself depend on the risk factors under investigation.^6^ Access to testing has also changed across the course of the pandemic,^7^ meaning earlier studies were more likely to focus on people with symptomatic disease or a history of travel, or on specific populations such as healthcare workers.

Serological population-based studies offer a different approach by testing members of a population uniformly, including people who might not be captured by routine testing. This approach not only reduces the risk of collider bias, but also can uncover previously undetected asymptomatic infections. Inclusion of asymptomatic SARS-CoV-2 infections in the analysis of risk factors is crucial, as asymptomatic individuals have been found to be as infectious as those with symptoms.^8^ Serology studies also offer the opportunity to identify determinants of anti-SARS-CoV-2 antibody titres, which are a recognised correlate of protection against future infection.^9,10^

The largest population-based serology studies done to date have explored several sociodemographic and clinical risk factors, but have not considered risk factors related to lifestyle, diet, or levels of physical activity.^3–5,11,12^ These studies have focused on IgG antibodies alone^11,12^ or relied on immunoassays with low sensitivity,^12^ potentially missing infections. They have also tended to be cross-sectional in design, so that reverse causality could potentially explain associations between symptomatic seropositivity and modifiable risk factors. Additionally, studies investigating determinants of antibody titres have focused on specific populations such as healthcare workers,^13,14^ limiting the generalisability of their findings.

We therefore undertook a prospective population-based study to uncover determinants of SARS-CoV-2 seropositivity and antibody titres, combining high statistical power with detailed assessment of sociodemographic, clinical, and behavioural risk factors, and supported by an assay with proven sensitivity for detection of SARS-CoV-2 antibodies in non-hospitalised adults with mild or moderate COVID-19.^15^

## Methods

### Study design and participants

COVIDENCE UK is a prospective, longitudinal, population-based observational study of COVID-19 in the UK population (www.qmul.ac.uk/covidence).^16^ Inclusion criteria were age 16 years or older and UK residence at enrolment, with no exclusion criteria. Participants were invited via a national media campaign to complete an online baseline questionnaire to capture: information on potential symptoms of COVID-19 experienced since Feb 1, 2020; results of any COVID-19 tests; and details of a wide range of potential risk factors for COVID-19 (appendix table S1). Online monthly follow-up questionnaires captured incident test-confirmed COVID-19 and symptoms of acute respiratory infection (appendix table S2). The study was launched on May 1, 2020.

The antibody study described here was introduced as an approved protocol amendment (amendment 3; November, 2020). Participants enrolled before the amendment were invited via email to participate in the antibody study and to give additional consent. As part of the antibody study, participants were invited to participate in serology testing from November, 2020. For this analysis, we included all participants who enrolled in the study between May 1 and November 2, 2020, partaking in serology testing who were not vaccinated against COVID-19 or who provided their dried blood spot sample on or before the date of their first COVID-19 vaccination. This paper reports findings from analysis of data collected up to April 18, 2021.

COVIDENCE UK was sponsored by Queen Mary University of London and approved by Leicester South Research Ethics Committee (ref 20/EM/0117). It is registered with ClinicalTrials.gov (NCT04330599).

### Procedures

Antibody study participants were sent a kit containing instructions, lancets, and blood spot collection cards, to be posted back to the study team. Once returned, the samples were logged by the study team and sent in batches to the Clinical Immunology Service at the Institute of Immunology and Immunotherapy of the University of Birmingham (Birmingham, UK). Up to two more test kits were offered to participants whose initial samples were found to be insufficient for testing. Blood spot samples were taken from November 6, 2020, to April 18, 2021.

Semi-quantitative determination of antibody titres in dried blood spot eluates was done using a commercially available ELISA that measures combined IgG, IgA, and IgM (IgGAM) responses to the SARS-CoV-2 trimeric spike glycoprotein (product code MK654; The Binding Site [TBS], Birmingham, UK). The SARS-CoV-2 spike used is a soluble, stabilised, trimeric glycoprotein truncated at the transmembrane region.^17,18^ This assay has been CE-marked with 98.3% (95% CI 96.4–99.4) specificity and 98.6% (92.6–100.0) sensitivity following RT-PCR-confirmed mild-to-moderate COVID-19 that did not result in hospitalisation.^15^ A cut-off ratio relative to the TBS cut-off calibrators was determined by plotting 624 pre-2019 negatives in a frequency histogram. A cut-off coefficient was then established for IgGAM (1.31), with ratio values classed as positive (≥1) or negative (<1). Dried blood spots were pre-diluted at a 1:40 dilution with 0.05% PBS-Tween using a Dynex Revelation automated absorbance microplate reader (Dynex Technologies; Chantilly, VA, USA). Plates were developed after 10 min using 3,3′,5,5′-tetramethylbenzidine core, and orthophosphoric acid used as a stop solution (both TBS). Optical densities at 450 nm were measured using the Dynex Revelation. Results of ELISA for detection of anti-S antibodies in dried blood spot eluates have previously been shown to have almost perfect agreement with those performed on serum (Cohen’s kappa = 0.83).^19^

### Outcomes

Study outcomes were presence versus absence of antibodies against SARS-CoV-2 (binary outcome assessed in all participants who did not report having tested positive for SARS-CoV-2 infection via RT-PCR or lateral flow test before enrolment), and antibody titres (continuous outcome measured in all seropositive participants).

### Independent variables

Eighty-eight putative risk factors for SARS-CoV-2 infection were selected *a priori*, covering sociodemographic, occupational, and lifestyle factors; longstanding medical conditions and prescribed medication use; Bacille Calmette Guérin and measles, mumps, and rubella vaccine status; and diet and supplemental micronutrient intake (appendix tables S1, S2). These factors, which were obtained from the baseline questionnaire, were included as independent variables in our models. To produce patient-level covariates for each class of medications investigated, participant responses were mapped to drug classes listed in the British National Formulary or the DrugBank and Electronic Medicines Compendium databases if not explicitly listed in the British National Formulary, as previously described.^16^ Index of Multiple Deprivation (IMD) 2019 scores were assigned according to participants’ postcodes, and categorised into quartiles. Duration of follow-up was defined as the number of days between the date of enrolment and the date of dried blood spot collection.

### Statistical analysis

Using the Stata powerlog program, we estimated that a minimum sample size of 10,964 would be required to detect a difference of at least 2% difference in the proportion of exposed vs. unexposed participants experiencing a given binary outcome [equivalent to an odds ratio (OR) of 1.08], with 90% power, for a binary exposure with maximum variability (probability 0.50 changing to 0.52) and a moderate correlation (R^2^=0.4) with other variables in a logistic regression model, using a two-sided test and 5% significance. The antibody study was a pragmatic study including all participants meeting the inclusion criteria, with no sample size specified.

Logistic regression models were used to estimate ORs and 95% CIs for potential determinants of SARS-CoV-2 seropositivity. Linear regression models with robust standard errors were used to estimate geometric mean ratios (GMRs) and 95% CIs for potential determinants of log-transformed antibody titres in seropositive participants. We first estimated ORs and GMRs in minimally adjusted models, and carried forward factors independently associated with each outcome at the 10% significance level to fully adjusted models. Both the minimally adjusted and fully adjusted models were controlled for age (<30 years, 30 to <40 years, 40 to <50 years, 50 to <60 years, 60 to <70 years, and ≥70 years), sex (male *vs* female), and duration of follow-up (days). We calculated p for trend for ordinal variables by re-running the regressions treating each ordinal variable in turn as continuous. Analyses were done for all participants with available data; missing data were not imputed. Correction for multiple comparisons was not applied, on the grounds that we were testing *a priori* hypotheses for all risk factors investigated.^20^

In a sensitivity analysis, we excluded participants from the seropositivity analysis who were classified as having had probable COVID-19 before enrolment on the basis of self-reported symptoms, using the symptom algorithm described and validated by Menni and colleagues.^21^

As antibody titres have been found to be associated with disease severity,^13,22^ we did an exploratory analysis to investigate the extent to which COVID-19 severity might explain associations between independent variables and antibody titres, by including this as an explanatory variable in the titre analysis. COVID-19 severity was classified into three groups: ‘asymptomatic’ (non-hospitalised seropositive participants, who either did not report any symptoms of acute respiratory infection or whose symptoms were classified as having <50% probability of being due to COVID-19, using the symptom algorithm by Menni and colleagues^21^); ‘symptomatic non-hospitalised’ (non-hospitalised seropositive participants who reported symptoms of acute respiratory infection that were classified as having ≥50% probability of being due to COVID-19, using the symptom algorithm^21^); and ‘hospitalised’ (seropositive participants who were hospitalised for treatment of COVID-19).

We present descriptive statistics as n (%), mean (SD), or median (IQR). Statistical analyses were done using Stata (version 14.2; StataCorp, College Station, TX, USA).

## Results

Serology data were available for 12,294 of the 15,853 participants who consented to participate in the antibody study. We excluded data from 1074 participants who had been vaccinated against SARS-CoV-2 before providing their dried blood spot sample (figure 1). Of the 11,220 participants included, 1774 (15.8%) tested positive for SARS-CoV-2 antibodies. For the analysis of determinants of seropositivity, we excluded 90 (0.8%) participants who reported a positive RT-PCR or lateral flow test result for SARS-CoV-2 infection before enrolment, leaving a sample size of 11,130 participants with 1696 seropositive cases (figure 1). Selected baseline characteristics of included participants are shown in table 1. 70.1% of participants were female, and 95.7% identified their ethnicity as White, with median age of 62.3 years (IQR 52.9–68.7; table 1).

**Figure 1:**
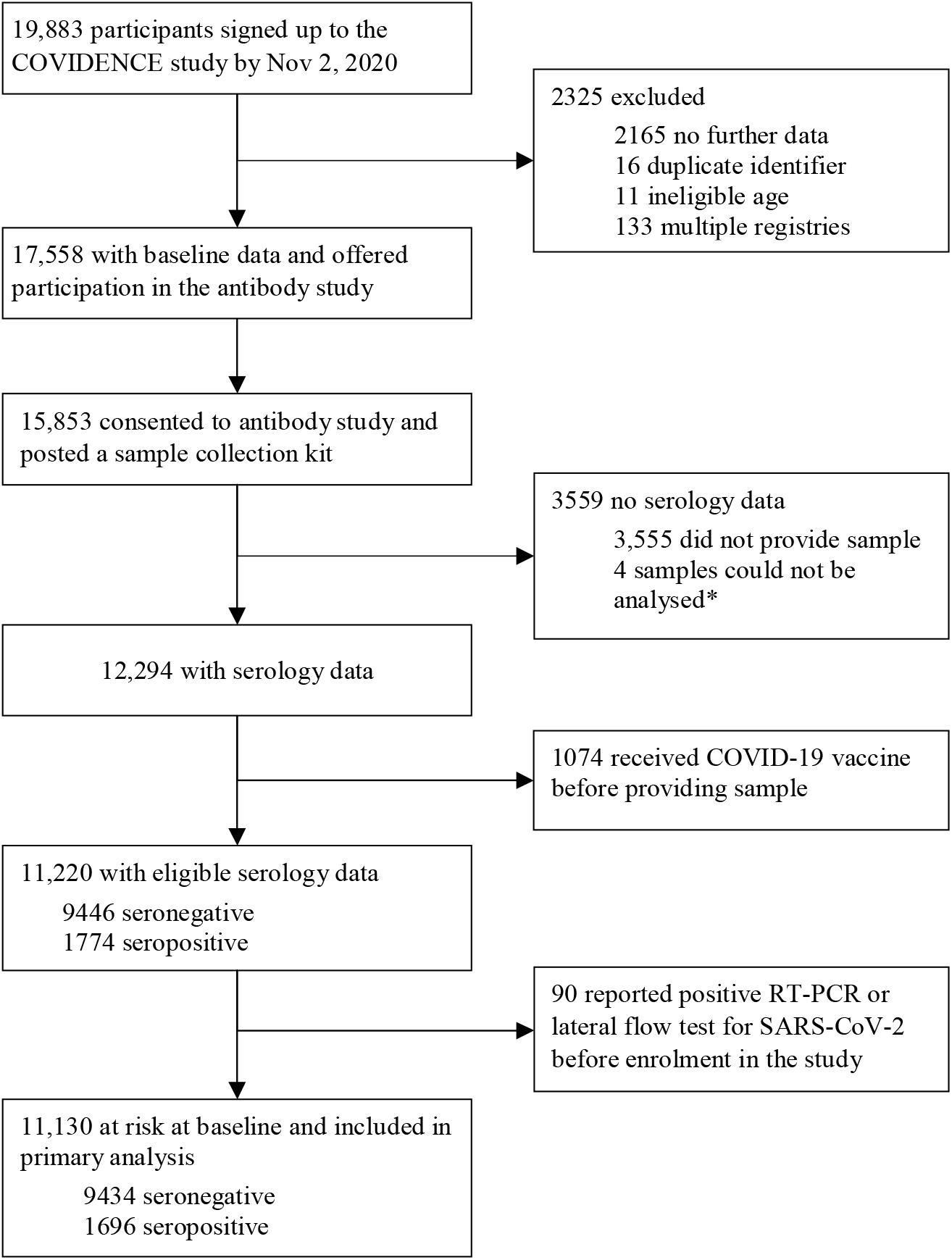
Study profile *103 participants provided insufficient samples, but 99 were successfully analysed upon repeat test.

**Table 1:**
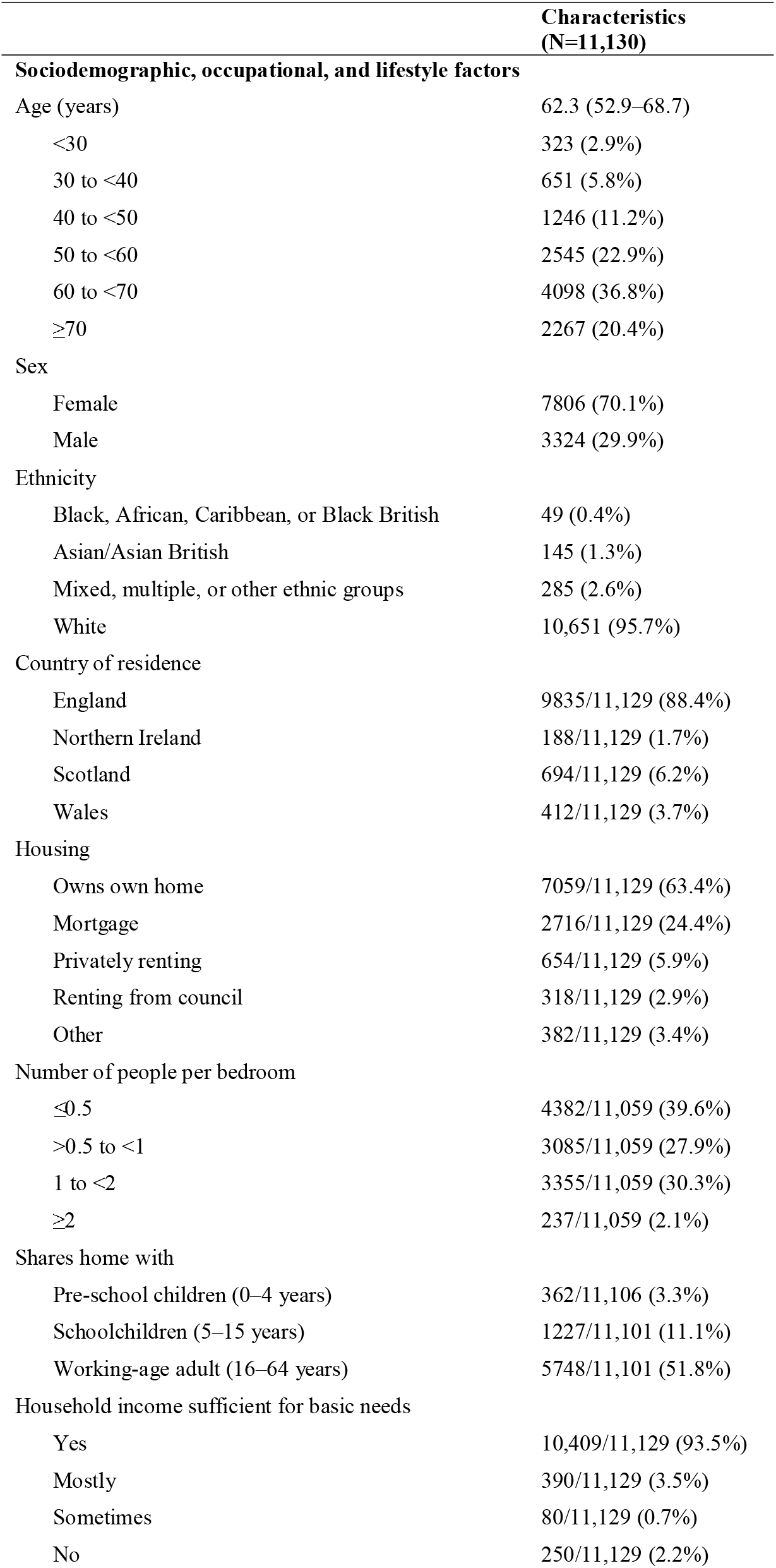
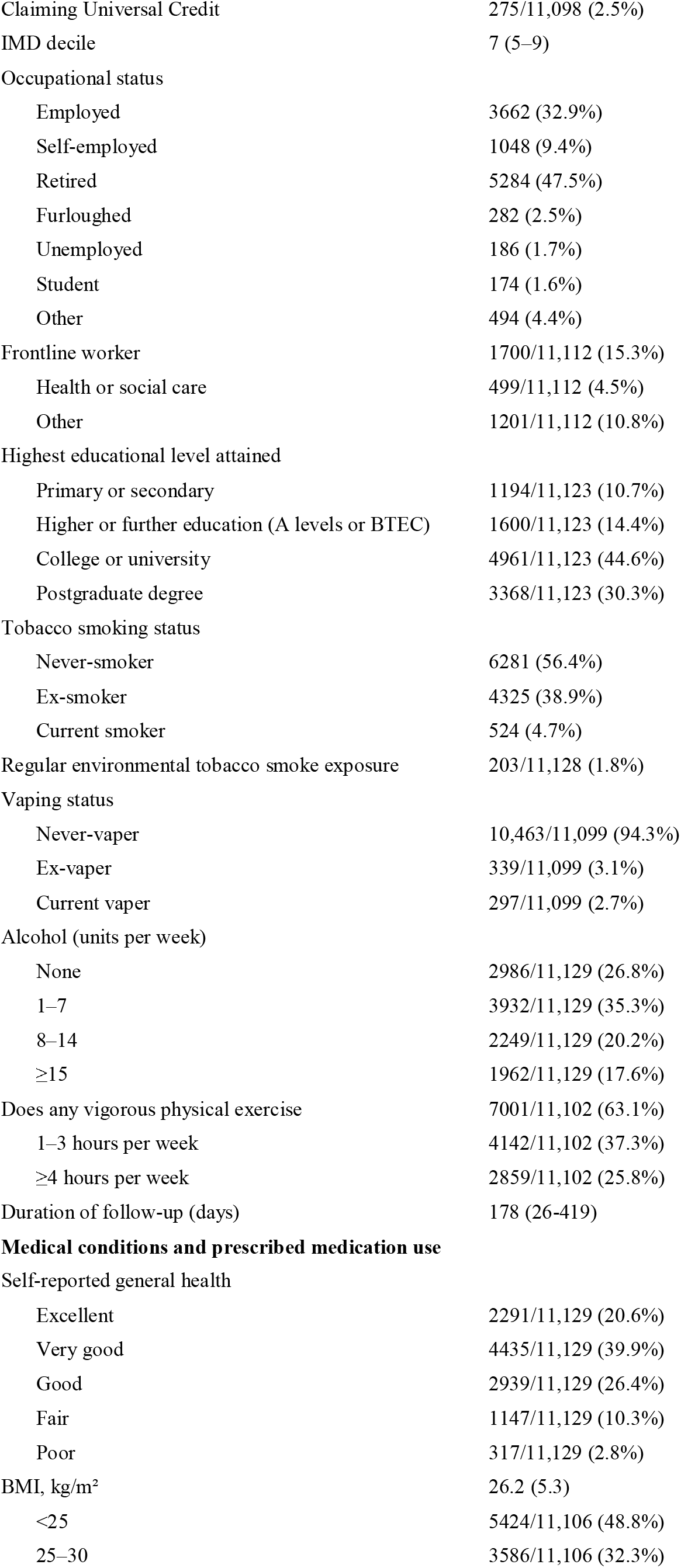
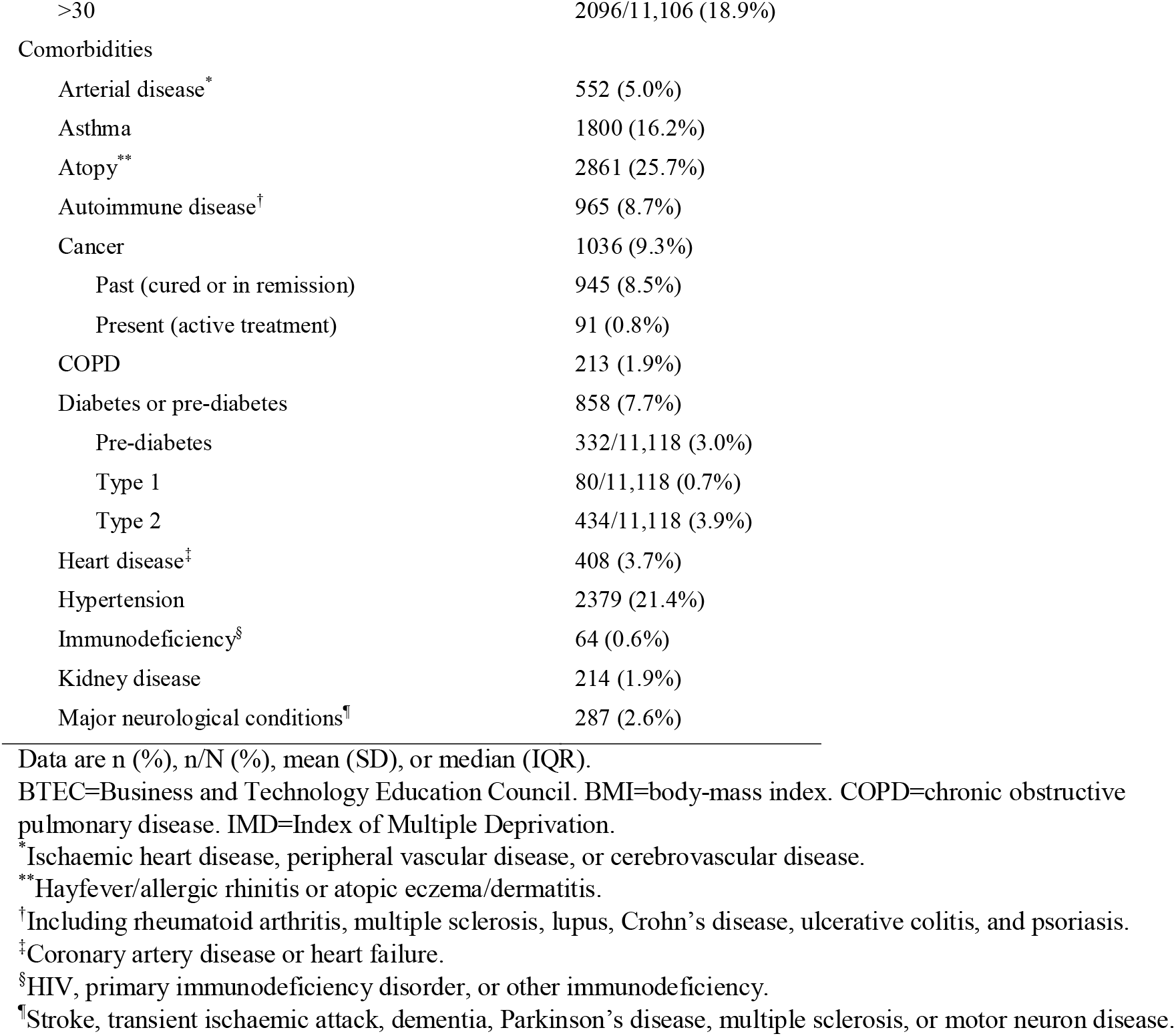
Baseline characteristics of participants included in seropositivity analysis

|  | Characteristics<br>(N=11,130) |
| --- | --- |
| **Sociodemographic, occupational, and lifestyle factors** |  |
| Age (years) | 62.3 (52.9–68.7) |
| <30 | 323 (2.9%) |
| 30 to <40 | 651 (5.8%) |
| 40 to <50 | 1246 (11.2%) |
| 50 to <60 | 2545 (22.9%) |
| 60 to <70 | 4098 (36.8%) |
| ≥70 | 2267 (20.4%) |
| Sex |  |
| Female | 7806 (70.1%) |
| Male | 3324 (29.9%) |
| Ethnicity |  |
| Black, African, Caribbean, or Black British | 49 (0.4%) |
| Asian/Asian British | 145 (1.3%) |
| Mixed, multiple, or other ethnic groups | 285 (2.6%) |
| White | 10,651 (95.7%) |
| Country of residence |  |
| England | 9835/11,129 (88.4%) |
| Northern Ireland | 188/11,129 (1.7%) |
| Scotland | 694/11,129 (6.2%) |
| Wales | 412/11,129 (3.7%) |
| Housing |  |
| Owns own home | 7059/11,129 (63.4%) |
| Mortgage | 2716/11,129 (24.4%) |
| Privately renting | 654/11,129 (5.9%) |
| Renting from council | 318/11,129 (2.9%) |
| Other | 382/11,129 (3.4%) |
| Number of people per bedroom |  |
| ≤0.5 | 4382/11,059 (39.6%) |
| >0.5 to <1 | 3085/11,059 (27.9%) |
| 1 to <2 | 3355/11,059 (30.3%) |
| ≥2 | 237/11,059 (2.1%) |
| Shares home with |  |
| Pre-school children (0–4 years) | 362/11,106 (3.3%) |
| Schoolchildren (5–15 years) | 1227/11,101 (11.1%) |
| Working-age adult (16–64 years) | 5748/11,101 (51.8%) |
| Household income sufficient for basic needs |  |
| Yes | 10,409/11,129 (93.5%) |
| Mostly | 390/11,129 (3.5%) |
| Sometimes | 80/11,129 (0.7%) |
| No | 250/11,129 (2.2%) |
| Claiming Universal Credit | 275/11,098 (2.5%) |
| IMD decile | 7 (5–9) |
| Occupational status |  |
| Employed | 3662 (32.9%) |
| Self-employed | 1048 (9.4%) |
| Retired | 5284 (47.5%) |
| Furloughed | 282 (2.5%) |
| Unemployed | 186 (1.7%) |
| Student | 174 (1.6%) |
| Other | 494 (4.4%) |
| Frontline worker | 1700/11,112 (15.3%) |
| Health or social care | 499/11,112 (4.5%) |
| Other | 1201/11,112 (10.8%) |
| Highest educational level attained |  |
| Primary or secondary | 1194/11,123 (10.7%) |
| Higher or further education (A levels or BTEC) | 1600/11,123 (14.4%) |
| College or university | 4961/11,123 (44.6%) |
| Postgraduate degree | 3368/11,123 (30.3%) |
| Tobacco smoking status |  |
| Never-smoker | 6281 (56.4%) |
| Ex-smoker | 4325 (38.9%) |
| Current smoker | 524 (4.7%) |
| Regular environmental tobacco smoke exposure | 203/11,128 (1.8%) |
| Vaping status |  |
| Never-vaper | 10,463/11,099 (94.3%) |
| Ex-vaper | 339/11,099 (3.1%) |
| Current vaper | 297/11,099 (2.7%) |
| Alcohol (units per week) |  |
| None | 2986/11,129 (26.8%) |
| 1–7 | 3932/11,129 (35.3%) |
| 8–14 | 2249/11,129 (20.2%) |
| ≥15 | 1962/11,129 (17.6%) |
| Does any vigorous physical exercise | 7001/11,102 (63.1%) |
| 1–3 hours per week | 4142/11,102 (37.3%) |
| ≥4 hours per week | 2859/11,102 (25.8%) |
| Duration of follow-up (days) | 178 (26–419) |
| **Medical conditions and prescribed medication use** |  |
| Self-reported general health |  |
| Excellent | 2291/11,129 (20.6%) |
| Very good | 4435/11,129 (39.9%) |
| Good | 2939/11,129 (26.4%) |
| Fair | 1147/11,129 (10.3%) |
| Poor | 317/11,129 (2.8%) |
| BMI, kg/m<sup>2</sup> | 26.2 (5.3) |
| <25 | 5424/11,106 (48.8%) |
| 25–30 | 3586/11,106 (32.3%) |
| >30 | 2096/11,106 (18.9%) |
| Comorbidities |  |
| Arterial disease<sup>*</sup> | 552 (5.0%) |
| Asthma | 1800 (16.2%) |
| Atopy<sup>**</sup> | 2861 (25.7%) |
| Autoimmune disease<sup>†</sup> | 965 (8.7%) |
| Cancer | 1036 (9.3%) |
| Past (cured or in remission) | 945 (8.5%) |
| Present (active treatment) | 91 (0.8%) |
| COPD | 213 (1.9%) |
| Diabetes or pre-diabetes | 858 (7.7%) |
| Pre-diabetes | 332/11,118 (3.0%) |
| Type 1 | 80/11,118 (0.7%) |
| Type 2 | 434/11,118 (3.9%) |
| Heart disease<sup>‡</sup> | 408 (3.7%) |
| Hypertension | 2379 (21.4%) |
| Immunodeficiency<sup>§</sup> | 64 (0.6%) |
| Kidney disease | 214 (1.9%) |
| Major neurological conditions<sup>¶</sup> | 287 (2.6%) |

Data are n (%), n/N (%), mean (SD), or median (IQR).
BTEC=Business and Technology Education Council. BMI=body-mass index. COPD=chronic obstructive pulmonary disease. IMD=Index of Multiple Deprivation.
<sup>\*</sup>Ischaemic heart disease, peripheral vascular disease, or cerebrovascular disease.
<sup>\*\*</sup>Hayfever/allergic rhinitis or atopic eczema/dermatitis.
<sup>†</sup>Including rheumatoid arthritis, multiple sclerosis, lupus, Crohn's disease, ulcerative colitis, and psoriasis.
<sup>‡</sup>Coronary artery disease or heart failure.
<sup>§</sup>HIV, primary immunodeficiency disorder, or other immunodeficiency.
<sup>¶</sup>Stroke, transient ischaemic attack, dementia, Parkinson's disease, multiple sclerosis, or motor neuron disease.

After adjustment for age, sex and duration of follow-up, 25 factors were independently associated with risk of SARS-CoV-2 seropositivity with p<0.10 (table 2). Appendix table S3 shows factors with no evidence of association. When the former factors were included together in a fully adjusted model, we observed that Asian/Asian British ethnicity (*vs* White), working as a frontline worker in a health or care setting (*vs* not working as a frontline worker), recent travel to a place of work or study, number of public transport journeys, visits to shops and other indoor public places, travel outside of the UK, high levels of alcohol consumption (≥15 units per week), high body-mass index (BMI; ≥25 kg/m^2^), sex hormone therapy (i.e. hormone replacement therapy and hormonal contraception), and use of vitamin D supplements were independently associated with increased risk of SARS-CoV-2 infection as indicated by antibody seropositivity (table 2). By contrast, postgraduate education (*vs* primary or secondary), passive smoking, high levels of light physical exercise (walking ≥10 h per week), and prescribed paracetamol use were independently associated with reduced risk of SARS-CoV-2 infection. In the fully adjusted model, the associations originally observed in minimally adjusted models for generational composition of households, living with a working-age adult, and lower impact physical activity no longer achieved conventional significance (table 2). Excluding the 796 participants with symptom-defined probable COVID-19, who did not have a positive PCR or lateral flow test result before enrolment, had little effect on our findings and associations with only 5 items were substantially attenuated in the minimally adjusted model, including environmental tobacco smoke exposure, public transport journeys, people per bedroom, dairy products intake, and use of sex hormone therapy (appendix table S4).

**Table 2:** Minimally adjusted and fully adjusted odds of seropositivity

|  | Seropositive participants<br>n/N (%) | Minimally adjusted model<sup>*</sup><br>OR (95% CI) | p value | Fully adjusted model<sup>†</sup><br>OR (95% CI) | p value |
| --- | --- | --- | --- | --- | --- |
| **Sociodemographic, occupational, and lifestyle factors** |  |  |  |  |  |
| Age (years) |  |  |  |  |  |
| <30 | 45/323 (13.9%) | 1.00 |  | 1.00 |  |
| 30 to <40 | 116/651 (17.8%) | 1.34 (0.92-1.95) | 0.13 | 1.42 (0.94-2.15) | 0.09 |
| 40 to <50 | 241/1246 (19.3%) | 1.48 (1.05-2.09) | 0.03 | 1.50 (1.01-2.24) | 0.04 |
| 50 to <60 | 419/2545 (16.5%) | 1.22 (0.88-1.71) | 0.23 | 1.31 (0.89-1.93) | 0.17 |
| 60 to <70 | 557/4098 (13.6%) | 0.98 (0.71-1.37) | 0.92 | 1.22 (0.82-1.82) | 0.34 |
| ≥70 | 318/2267 (14.0%) | 1.03 (0.73-1.44) | 0.89 | 1.41 (0.92-2.15) | 0.11 |
| *p for trend* | .. | .. | 0.89 | .. | 0.91 |
| Sex |  |  |  |  |  |
| Female | 1177/7806 (15.1%) | 1.00 |  | 1.00 |  |
| Male | 519/3324 (15.6%) | 1.10 (0.98-1.24) | 0.10 | 1.06 (0.93-1.20) | 0.39 |
| Ethnicity |  |  |  |  |  |
| White | 1598/10,651 (15.0%) | 1.00 |  | 1.00 |  |
| Black, African, Caribbean, or Black British | 10/49 (20.4%) | 1.30 (0.96-1.76) | 0.09 | 1.24 (0.90-1.71) | 0.20 |
| Asian/Asian British | 34/145 (23.4%) | 1.65 (1.12-2.45) | 0.01 | 1.65 (1.10-2.49) | 0.02 |
| Mixed, multiple, or other ethnic groups | 54/285 (18.9%) | 1.36 (0.67-2.73) | 0.40 | 1.14 (0.52-2.50) | 0.74 |
| Housing |  |  |  |  |  |
| Owns own home | 991/7059 (14.0%) | 1.00 |  | 1.00 |  |
| Mortgage | 481/2716 (17.7%) | 1.14 (0.99-1.32) | 0.07 | 1.09 (0.93-1.27) | 0.28 |
| Privately renting | 108/654 (16.5%) | 1.12 (0.88-1.42) | 0.36 | 1.23 (0.95-1.58) | 0.11 |
| Renting from council | 56/318 (17.6%) | 1.20 (0.89-1.62) | 0.24 | 1.32 (0.94-1.87) | 0.11 |
| Other | 60/382 (15.7%) | 1.09 (0.79-1.50) | 0.60 | 1.11 (0.80-1.55) | 0.53 |
| Claiming Universal Credit |  |  |  |  |  |
| No | 1654/10,823 (15.3%) | 1.00 |  | 1.00 |  |
| Yes | 35/275 (12.7%) | 0.73 (0.51-1.05) | 0.09 | 0.72 (0.49-1.05) | 0.09 |
| Number of people per bedroom |  |  |  |  |  |
| ≤0.5 | 590/4382 (13.5%) | 1.00 |  | 1.00 |  |
| >0.5 to <1 | 499/3085 (16.2%) | 1.18 (1.04-1.35) | 0.01 | 1.04 (0.90-1.21) | 0.57 |
| 1 to <2 | 559/3355 (16.7%) | 1.13 (0.98-1.30) | 0.09 | 0.97 (0.83-1.14) | 0.73 |
| ≥2 | 35/237 (14.8%) | 0.96 (0.66-1.41) | 0.85 | 0.73 (0.48-1.11) | 0.15 |
| *p for trend* | .. | .. | 0.13 | .. | 0.42 |
| Multigenerational household |  |  |  |  |  |
| Living alone | 265/1997 (13.3%) | 1.00 |  | 1.00 |  |
| Single generation | 925/6055 (15.3%) | 1.17 (1.01-1.36) | 0.03 | 1.14 (0.95-1.38) | 0.17 |
| Two-generation | 488/2990 (16.3%) | 1.19 (1.01-1.40) | 0.04 | 1.17 (0.94-1.45) | 0.15 |
| Three-generation | 18/88 (20.5%) | 1.61 (0.94-2.76) | 0.08 | 1.64 (0.91-2.96) | 0.10 |
| *p for trend* | .. | .. | 0.03 | .. | 0.08 |
| Shares home with working-age adult (16–64 years) |  |  |  |  |  |
| No | 724/5353 (13.5%) | 1.00 |  | 1.00 |  |
| Yes | 968/5748 (16.8%) | 1.21 (1.06-1.37) | 0.003 | 1.08 (0.93-1.26) | 0.32 |
| Highest educational level attained |  |  |  |  |  |
| Primary or secondary | 198 (16.6%) | 1.00 |  | 1.00 |  |
| Higher or further education (A levels or BTEC) | 244 (15.3%) | 0.92 (0.75-1.13) | 0.42 | 0.94 (0.76-1.17) | 0.60 |
| College or university | 777 (15.7%) | 0.95 (0.80-1.13) | 0.54 | 0.94 (0.78-1.12) | 0.47 |
| Postgraduate degree | 476 (14.1%) | 0.83 (0.69-0.99) | 0.04 | 0.82 (0.67-0.99) | 0.04 |
| *p for trend* | .. | .. | 0.04 | .. | 0.03 |
| Frontline worker |  |  |  |  |  |
| No | 1369/9422 (14.5%) | 1.00 |  | 1.00 |  |
| Non-health | 197/1201 (16.4%) | 1.13 (0.95-1.33) | 0.17 | 1.02 (0.85-1.22) | 0.86 |
| Health or care | 130/499 (26.1%) | 2.02 (1.63-2.50) | <0.001 | 1.86 (1.48-2.33) | <0.001 |
| Travel to place of work or study in past week |  |  |  |  |  |
| No | 669/5031 (13.3%) | 1.00 |  | 1.00 |  |
| Yes | 1014/5981 (17.0%) | 1.31 (1.17-1.46) | <0.001 | 1.20 (1.07-1.35) | <0.001 |
| Number of public transport journeys per week |  |  |  |  |  |
| 0 | 1502/9923 (15.1) | 1.00 |  | 1.00 |  |
| 1-5 | 135/849 (15.9) | 1.24 (1.02-1.51) | 0.03 | 1.19 (0.97-1.47) | 0.09 |
| ≥6 | 55/322 (17.1) | 1.32 (0.98-1.78) | 0.07 | 1.24 (0.90-1.69) | 0.18 |
| *p for trend* | .. | .. | 0.01 | .. | 0.048 |
| Number of visits to shops and other indoor public places per week |  |  |  |  |  |
| 0 | 220/1514 (14.5) | 1.00 |  | 1.00 |  |
| 1-2 | 560/3611 (15.5) | 1.15 (0.97-1.37) | 0.10 | 1.12 (0.94-1.34) | 0.20 |
| 2-4 | 413/2578 (16.0) | 1.33 (1.11-1.60) | 0.002 | 1.27 (1.05-1.53) | 0.02 |
| ≥5 | 503/3413 (14.7) | 1.36 (1.13-1.64) | 0.001 | 1.29 (1.06-1.57) | 0.01 |
| *p for trend* | .. | .. | <0.001 | .. | 0.01 |
| Travel outside of the UK between November 2019, and February 2021<sup>‡</sup> |  |  |  |  |  |
| No | 926/6529 (14.2%) | 1.00 |  | 1.00 |  |
| Yes | 572/3507 (16.3%) | 1.19 (1.07-1.34) | 0.002 | 1.20 (1.07-1.36) | 0.002 |
| Tobacco smoking status |  |  |  |  |  |
| Never-smoker | 930/6281 (14.8%) | 1.00 |  | 1.00 |  |
| Ex-smoker | 690/4325 (16.0%) | 1.10 (0.98-1.22) | 0.09 | 1.05 (0.94-1.18) | 0.42 |
| Current smoker | 76/524 (14.5%) | 0.90 (0.70-1.16) | 0.43 | 0.81 (0.61-1.07) | 0.14 |
| Regular environmental tobacco smoke exposure |  |  |  |  |  |
| No | 1672/10,925 (15.3%) | 1.00 |  | 1.00 |  |
| Yes | 23/203 (11.3%) | 0.65 (0.42-1.01) | 0.05 | 0.59 (0.37-0.95) | 0.03 |
| Alcohol (units per week) |  |  |  |  |  |
| None | 438/2986 (14.7%) | 1.00 |  | 1.00 |  |
| 1-7 | 610/3932 (15.5%) | 1.11 (0.97-1.27) | 0.13 | 1.09 (0.95-1.26) | 0.22 |
| 8-14 | 318/2249 (14.1%) | 1.00 (0.86-1.17) | 0.99 | 1.01 (0.85-1.19) | 0.92 |
| ≥15 | 330/1962 (16.8%) | 1.22 (1.04-1.43) | 0.02 | 1.23 (1.04-1.46) | 0.02 |
| *p for trend* | .. | .. | 0.07 | .. | 0.06 |
| Light physical exercise (hours per week) |  |  |  |  |  |
| 0-4 | 614/3631 (16.9%) | 1.00 |  | 1.00 |  |
| 5-9 | 613/3714 (16.5%) | 1.01 (0.90-1.15) | 0.83 | 1.03 (0.90-1.17) | 0.69 |
| ≥10 | 468/3764 (12.4%) | 0.78 (0.68-0.89) | 0.000 | 0.80 (0.70-0.93) | 0.003 |
| *p for trend* | .. | .. | <0.001 | .. | 0.003 |
| Lower impact physical activity (hours per week) |  |  |  |  |  |
| 0 | 991/6272 (15.8%) | 1.00 |  | 1.00 |  |
| 1 | 322/2131 (15.1%) | 0.95 (0.83-1.09) | 0.46 | 0.97 (0.84-1.12) | 0.71 |
| ≥2 | 378/2695 (14.0%) | 0.87 (0.77-1.00) | 0.04 | 0.93 (0.81-1.07) | 0.33 |
| *p for trend* | .. | .. | 0.04 | .. | 0.33 |
| **Diet and supplemental micronutrient intake** |  |  |  |  |  |
| Portions of dairy products or calcium-fortified alternatives per day |  |  |  |  |  |
| 0–1 | 474/2928 (16.2%) | 1.00 |  | 1.00 |  |
| 2 | 460/3254 (14.1%) | 0.86 (0.75-0.99) | 0.04 | 0.88 (0.76-1.02) | 0.08 |
| 3–5 | 389/2650 (14.7%) | 0.90 (0.78-1.04) | 0.17 | 0.92 (0.79-1.07) | 0.29 |
| ≥6 | 368/2269 (16.2%) | 1.03 (0.88-1.20) | 0.71 | 1.04 (0.89-1.22) | 0.63 |
| *p for trend* | .. | .. | 0.71 | .. | 0.62 |
| **Medical conditions and prescribed medication use** |  |  |  |  |  |
| BMI, kg/m<sup>2</sup> |  |  |  |  |  |
| <25 | 741/5424 (13.7%) | 1.00 |  | 1.00 |  |
| 25–30 | 604/3586 (16.8%) | 1.27 (1.13-1.43) | <0.001 | 1.25 (1.10-1.41) | <0.001 |
| >30 | 347/2096 (16.6%) | 1.19 (1.03-1.37) | 0.01 | 1.23 (1.06-1.43) | 0.01 |
| *p for trend* | .. | .. | 0.002 | .. | 0.001 |
| COPD |  |  |  |  |  |
| No | 1656/10,917 (15.2%) | 1.00 |  | 1.00 |  |
| Yes | 40/213 (18.8%) | 1.36 (0.95-1.93) | 0.09 | 1.41 (0.98-2.04) | 0.07 |
| Paracetamol |  |  |  |  |  |
| No | 1643/10,691 (15.4%) | 1.00 |  | 1.00 |  |
| Yes | 53/439 (12.1%) | 0.75 (0.56-1.01) | 0.06 | 0.71 (0.53-0.97) | 0.03 |
| Sex hormone therapy |  |  |  |  |  |
| No | 1551 (15.1%) | 1.00 |  | 1.00 |  |
| Yes | 145 (17.4%) | 1.18 (0.97-1.43) | 0.10 | 1.25 (1.02-1.52) | 0.03 |
| Vitamin D (over the counter or prescribed) |  |  |  |  |  |
| No | 1066/7300 (14.6%) | 1.00 |  | 1.00 |  |
| Yes | 630/3830 (16.4%) | 1.10 (0.99-1.23) | 0.08 | 1.16 (1.03-1.30) | 0.01 |
| Follow-up duration (days) | .. | 1.003 (1.002-1.004) | <0.001 | 1.003 (1.003-1.004) | <0.001 |

Descriptive data are n/N (%) indicating number of seropositive participants (n) and total per category (N). BMI=body-mass index. BTEC=Business and Technology Education Council. COPD=chronic obstructive pulmonary disease. OR=odds ratio.
\*Adjusted for age, sex, and duration of follow-up.
†Adjusted for all factors shown and duration of follow-up. The fully adjusted analysis includes 10,734 participants with data available for all factors.
‡The 1094 participants with unknown or missing travel status were included in the analysis as a separate category.

When investigating associations with antibody titres, analysed as a continuous outcome in the subset of seropositive participants only, we found that 35 factors were independently associated with antibody titres with p<0.10 after adjustment for age, sex, and duration of follow-up (table 3). The distribution of titres for three of these factors—ethnicity, frontline worker status, and COVID-19 severity—are shown in figure 2, with higher medians for non-White ethnicities, health or social care frontline workers, and participants who were hospitalised for treatment of COVID-19. Appendix table S5 shows factors with no evidence of association with antibody titre. When the 33 factors were included together in a fully adjusted model, we found that Asian/Asian British ethnicity (*vs* White), having a mortgage (*vs* owning own home), working as a frontline worker in a health or care setting, being an ex-smoker (*vs* a never-smoker), visits to shops and other indoor public places, travel outside of the UK, taking multivitamin supplements, consuming at least two portions of dairy products or calcium-fortified alternatives (*vs* 0–1 portions), and high BMI were associated with higher antibody titres, whereas high levels of fruit, vegetable, or salad consumption and reporting feeling anxious or depressed at baseline were associated with lower antibody titres (table 3). p-for-trend analyses suggested higher antibody titres with increasing intake of dairy or calcium-fortified alternatives and increasing BMI and lower antibody titres with increasing fruit, vegetable, or salad consumption (table 3). The associations in minimally adjusted models with chronic obstructive pulmonary disease, poor self-reported general health, and use of metformin or statins were attenuated in the fully adjusted model and were no longer statistically significant (table 3).

**Table 3:**
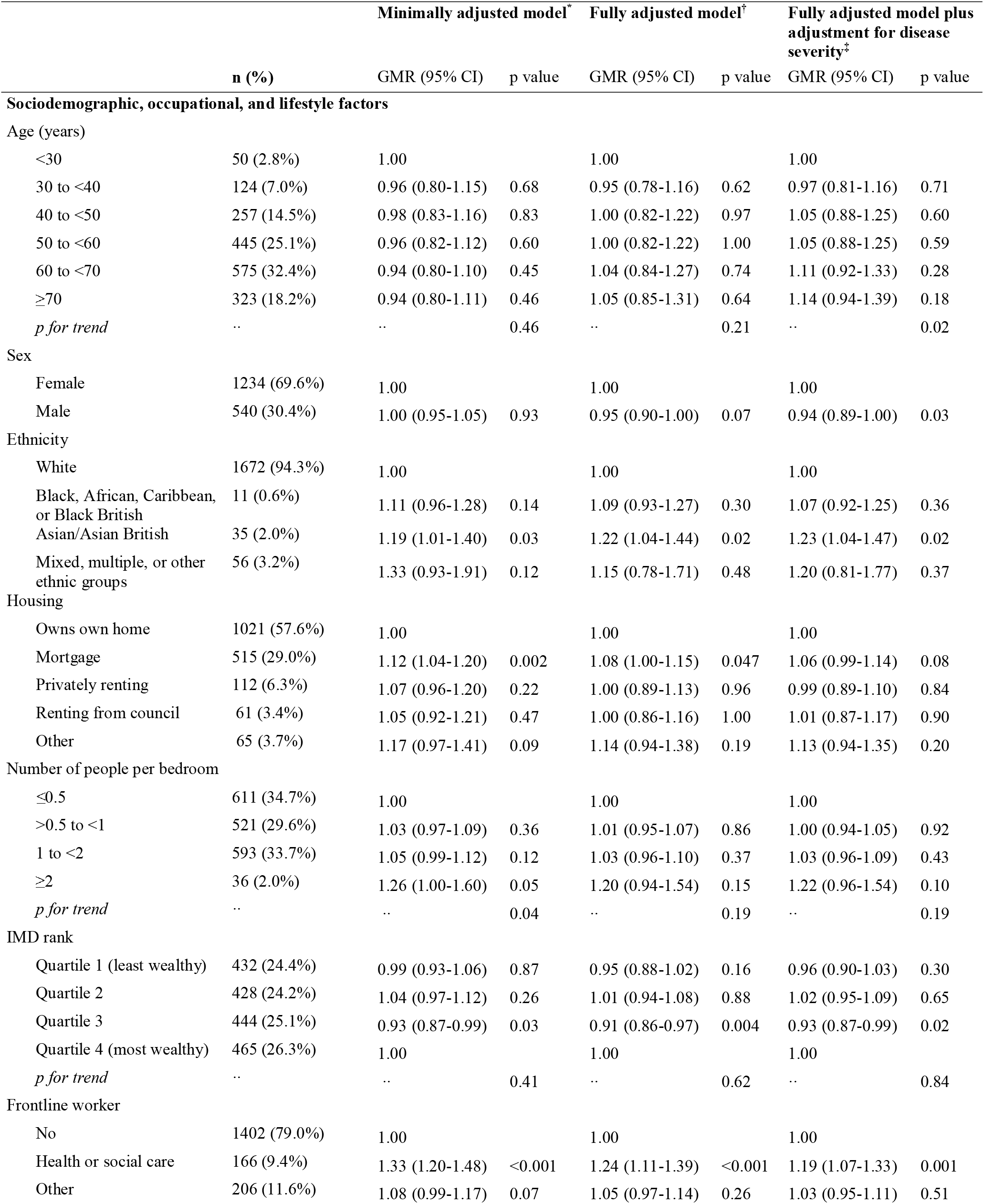
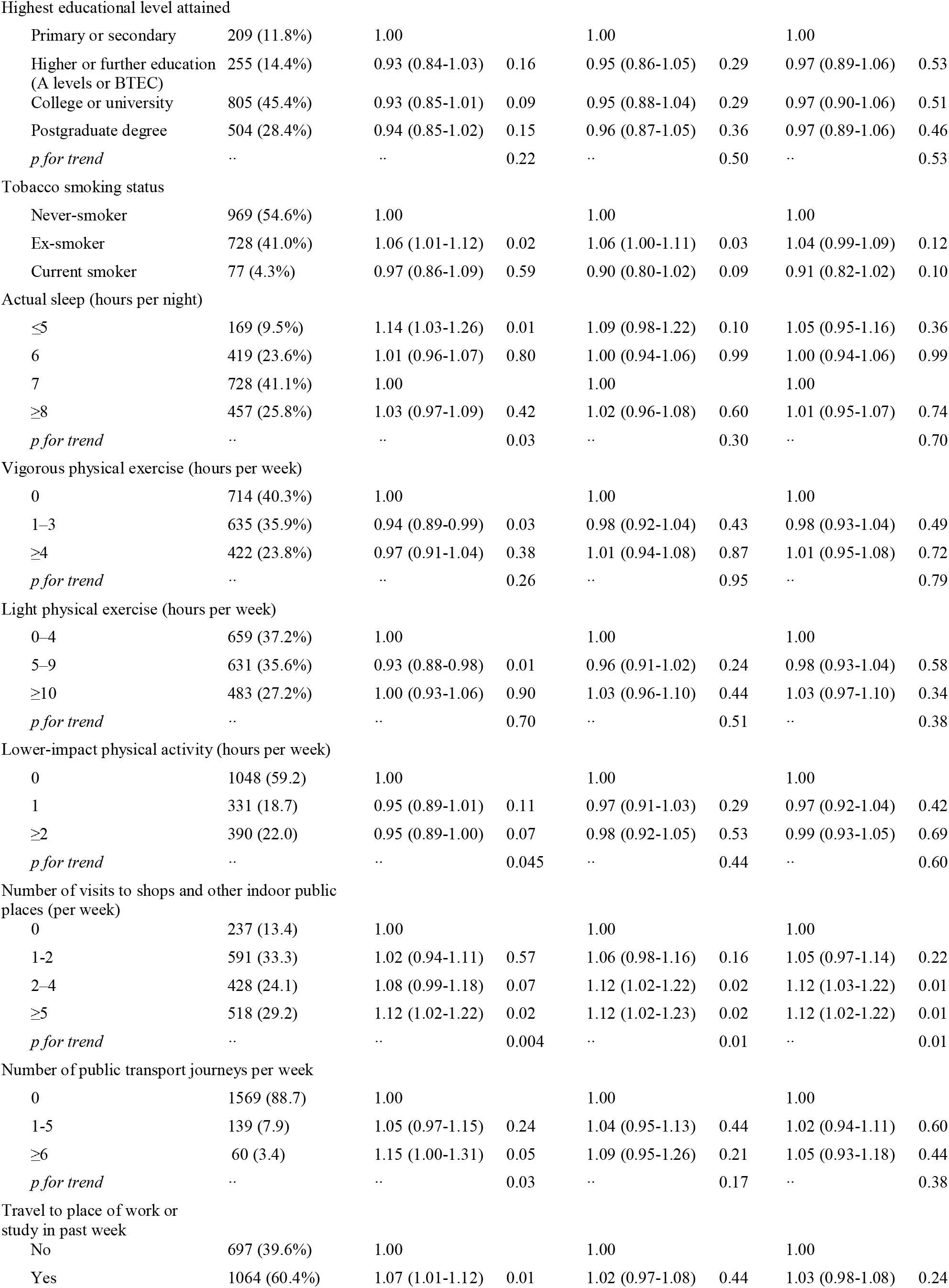
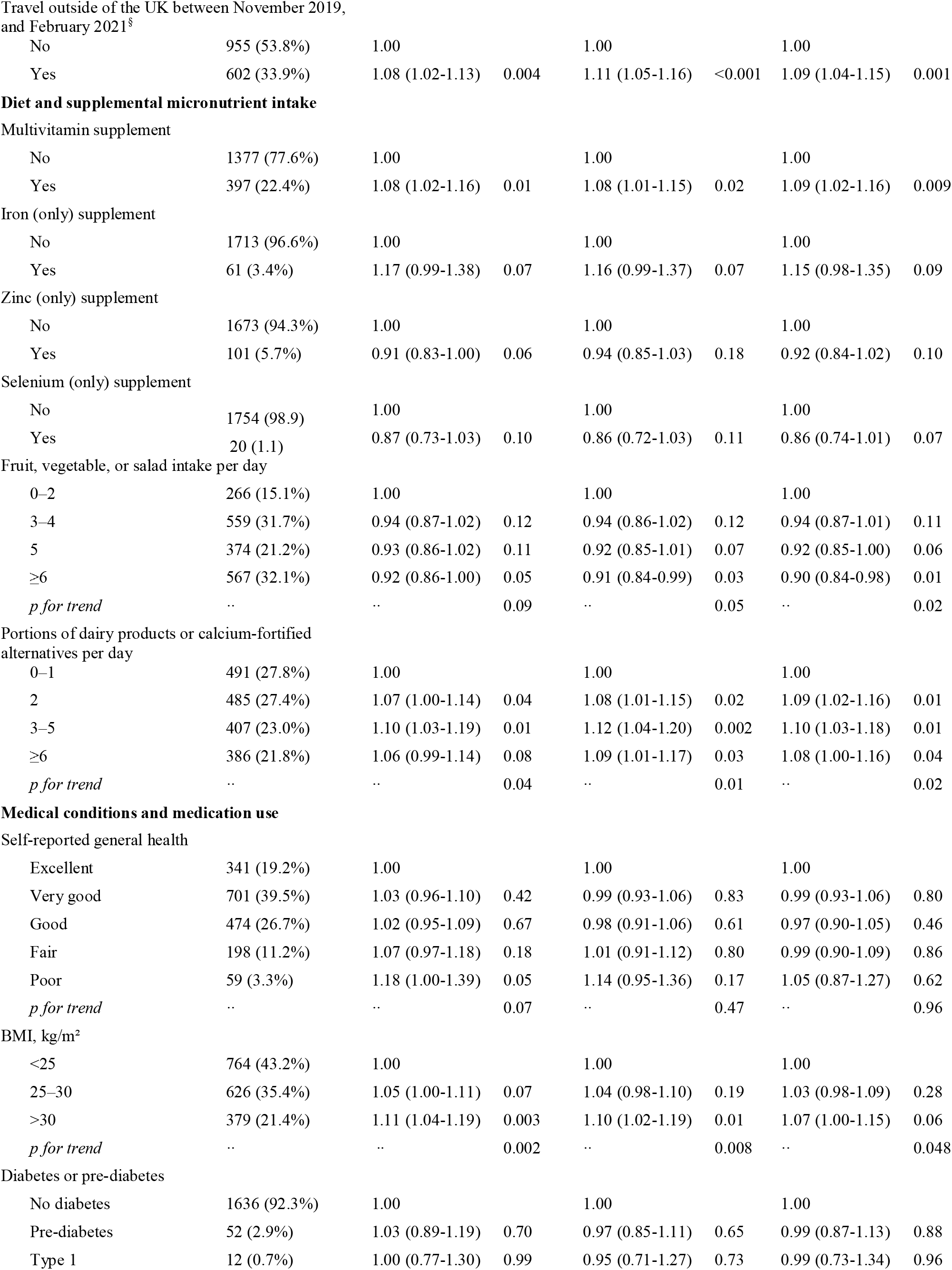
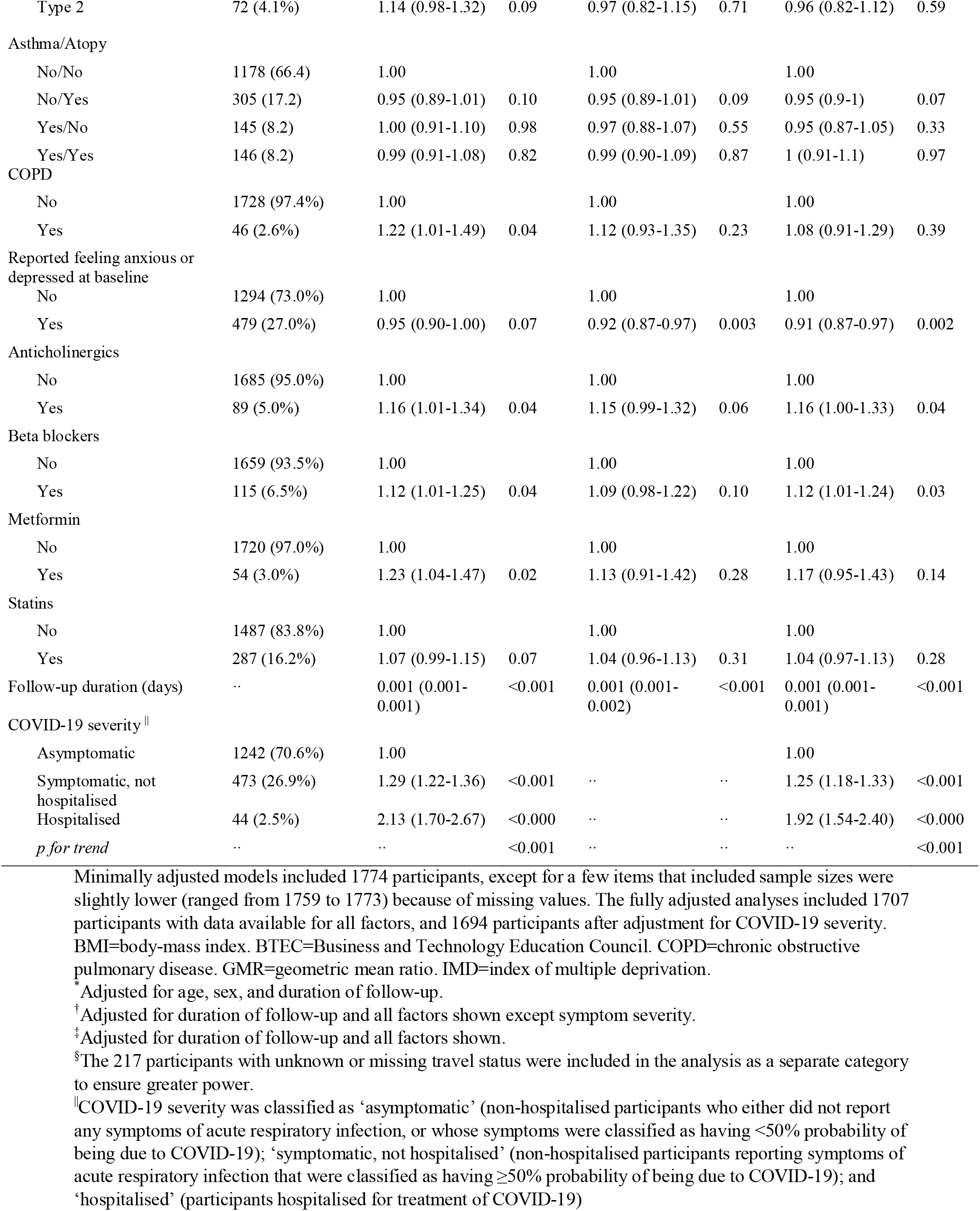
Minimally adjusted and fully adjusted geometric mean ratios of antibody titres in seropositive participants, with exploratory analysis of disease severity

|  |  | Minimally adjusted model* |  | Fully adjusted model† |  | Fully adjusted model plus adjustment for disease severity‡ |  |
| --- | --- | --- | --- | --- | --- | --- | --- |
|  | n (%) | GMR (95% CI) | p value | GMR (95% CI) | p value | GMR (95% CI) | p value |
| Sociodemographic, occupational, and lifestyle factors |  |  |  |  |  |  |  |
| Age (years) |  |  |  |  |  |  |  |
| <30 | 50 (2.8%) | 1.00 |  | 1.00 |  | 1.00 |  |
| 30 to <40 | 124 (7.0%) | 0.96 (0.80-1.15) | 0.68 | 0.95 (0.78-1.16) | 0.62 | 0.97 (0.81-1.16) | 0.71 |
| 40 to <50 | 257 (14.5%) | 0.98 (0.83-1.16) | 0.83 | 1.00 (0.82-1.22) | 0.97 | 1.05 (0.88-1.25) | 0.60 |
| 50 to <60 | 445 (25.1%) | 0.96 (0.82-1.12) | 0.60 | 1.00 (0.82-1.22) | 1.00 | 1.05 (0.88-1.25) | 0.59 |
| 60 to <70 | 575 (32.4%) | 0.94 (0.80-1.10) | 0.45 | 1.04 (0.84-1.27) | 0.74 | 1.11 (0.92-1.33) | 0.28 |
| ≥70 | 323 (18.2%) | 0.94 (0.80-1.11) | 0.46 | 1.05 (0.85-1.31) | 0.64 | 1.14 (0.94-1.39) | 0.18 |
| p for trend | .. | .. | 0.46 | .. | 0.21 | .. | 0.02 |
| Sex |  |  |  |  |  |  |  |
| Female | 1234 (69.6%) | 1.00 |  | 1.00 |  | 1.00 |  |
| Male | 540 (30.4%) | 1.00 (0.95-1.05) | 0.93 | 0.95 (0.90-1.00) | 0.07 | 0.94 (0.89-1.00) | 0.03 |
| Ethnicity |  |  |  |  |  |  |  |
| White | 1672 (94.3%) | 1.00 |  | 1.00 |  | 1.00 |  |
| Black, African, Caribbean, or Black British | 11 (0.6%) | 1.11 (0.96-1.28) | 0.14 | 1.09 (0.93-1.27) | 0.30 | 1.07 (0.92-1.25) | 0.36 |
| Asian/Asian British | 35 (2.0%) | 1.19 (1.01-1.40) | 0.03 | 1.22 (1.04-1.44) | 0.02 | 1.23 (1.04-1.47) | 0.02 |
| Mixed, multiple, or other ethnic groups | 56 (3.2%) | 1.33 (0.93-1.91) | 0.12 | 1.15 (0.78-1.71) | 0.48 | 1.20 (0.81-1.77) | 0.37 |
| Housing |  |  |  |  |  |  |  |
| Owns own home | 1021 (57.6%) | 1.00 |  | 1.00 |  | 1.00 |  |
| Mortgage | 515 (29.0%) | 1.12 (1.04-1.20) | 0.002 | 1.08 (1.00-1.15) | 0.047 | 1.06 (0.99-1.14) | 0.08 |
| Privately renting | 112 (6.3%) | 1.07 (0.96-1.20) | 0.22 | 1.00 (0.89-1.13) | 0.96 | 0.99 (0.89-1.10) | 0.84 |
| Renting from council | 61 (3.4%) | 1.05 (0.92-1.21) | 0.47 | 1.00 (0.86-1.16) | 1.00 | 1.01 (0.87-1.17) | 0.90 |
| Other | 65 (3.7%) | 1.17 (0.97-1.41) | 0.09 | 1.14 (0.94-1.38) | 0.19 | 1.13 (0.94-1.35) | 0.20 |
| Number of people per bedroom |  |  |  |  |  |  |  |
| ≤0.5 | 611 (34.7%) | 1.00 |  | 1.00 |  | 1.00 |  |
| >0.5 to <1 | 521 (29.6%) | 1.03 (0.97-1.09) | 0.36 | 1.01 (0.95-1.07) | 0.86 | 1.00 (0.94-1.05) | 0.92 |
| 1 to <2 | 593 (33.7%) | 1.05 (0.99-1.12) | 0.12 | 1.03 (0.96-1.10) | 0.37 | 1.03 (0.96-1.09) | 0.43 |
| ≥2 | 36 (2.0%) | 1.26 (1.00-1.60) | 0.05 | 1.20 (0.94-1.54) | 0.15 | 1.22 (0.96-1.54) | 0.10 |
| p for trend | .. | .. | 0.04 | .. | 0.19 | .. | 0.19 |
| IMD rank |  |  |  |  |  |  |  |
| Quartile 1 (least wealthy) | 432 (24.4%) | 0.99 (0.93-1.06) | 0.87 | 0.95 (0.88-1.02) | 0.16 | 0.96 (0.90-1.03) | 0.30 |
| Quartile 2 | 428 (24.2%) | 1.04 (0.97-1.12) | 0.26 | 1.01 (0.94-1.08) | 0.88 | 1.02 (0.95-1.09) | 0.65 |
| Quartile 3 | 444 (25.1%) | 0.93 (0.87-0.99) | 0.03 | 0.91 (0.86-0.97) | 0.004 | 0.93 (0.87-0.99) | 0.02 |
| Quartile 4 (most wealthy) | 465 (26.3%) | 1.00 |  | 1.00 |  | 1.00 |  |
| p for trend | .. | .. | 0.41 | .. | 0.62 | .. | 0.84 |
| Frontline worker |  |  |  |  |  |  |  |
| No | 1402 (79.0%) | 1.00 |  | 1.00 |  | 1.00 |  |
| Health or social care | 166 (9.4%) | 1.33 (1.20-1.48) | <0.001 | 1.24 (1.11-1.39) | <0.001 | 1.19 (1.07-1.33) | 0.001 |
| Other | 206 (11.6%) | 1.08 (0.99-1.17) | 0.07 | 1.05 (0.97-1.14) | 0.26 | 1.03 (0.95-1.11) | 0.51 |
| Highest educational level attained |  |  |  |  |  |  |  |
| Primary or secondary | 209 (11.8%) | 1.00 |  | 1.00 |  | 1.00 |  |
| Higher or further education<br>(A levels or BTEC) | 255 (14.4%) | 0.93 (0.84-1.03) | 0.16 | 0.95 (0.86-1.05) | 0.29 | 0.97 (0.89-1.06) | 0.53 |
| College or university | 805 (45.4%) | 0.93 (0.85-1.01) | 0.09 | 0.95 (0.88-1.04) | 0.29 | 0.97 (0.90-1.06) | 0.51 |
| Postgraduate degree | 504 (28.4%) | 0.94 (0.85-1.02) | 0.15 | 0.96 (0.87-1.05) | 0.36 | 0.97 (0.89-1.06) | 0.46 |
| *p for trend* | .. | .. | 0.22 | .. | 0.50 | .. | 0.53 |
| Tobacco smoking status |  |  |  |  |  |  |  |
| Never-smoker | 969 (54.6%) | 1.00 |  | 1.00 |  | 1.00 |  |
| Ex-smoker | 728 (41.0%) | 1.06 (1.01-1.12) | 0.02 | 1.06 (1.00-1.11) | 0.03 | 1.04 (0.99-1.09) | 0.12 |
| Current smoker | 77 (4.3%) | 0.97 (0.86-1.09) | 0.59 | 0.90 (0.80-1.02) | 0.09 | 0.91 (0.82-1.02) | 0.10 |
| Actual sleep (hours per night) |  |  |  |  |  |  |  |
| ≤5 | 169 (9.5%) | 1.14 (1.03-1.26) | 0.01 | 1.09 (0.98-1.22) | 0.10 | 1.05 (0.95-1.16) | 0.36 |
| 6 | 419 (23.6%) | 1.01 (0.96-1.07) | 0.80 | 1.00 (0.94-1.06) | 0.99 | 1.00 (0.94-1.06) | 0.99 |
| 7 | 728 (41.1%) | 1.00 |  | 1.00 |  | 1.00 |  |
| ≥8 | 457 (25.8%) | 1.03 (0.97-1.09) | 0.42 | 1.02 (0.96-1.08) | 0.60 | 1.01 (0.95-1.07) | 0.74 |
| *p for trend* | .. | .. | 0.03 | .. | 0.30 | .. | 0.70 |
| Vigorous physical exercise (hours per week) |  |  |  |  |  |  |  |
| 0 | 714 (40.3%) | 1.00 |  | 1.00 |  | 1.00 |  |
| 1–3 | 635 (35.9%) | 0.94 (0.89-0.99) | 0.03 | 0.98 (0.92-1.04) | 0.43 | 0.98 (0.93-1.04) | 0.49 |
| ≥4 | 422 (23.8%) | 0.97 (0.91-1.04) | 0.38 | 1.01 (0.94-1.08) | 0.87 | 1.01 (0.95-1.08) | 0.72 |
| *p for trend* | .. | .. | 0.26 | .. | 0.95 | .. | 0.79 |
| Light physical exercise (hours per week) |  |  |  |  |  |  |  |
| 0–4 | 659 (37.2%) | 1.00 |  | 1.00 |  | 1.00 |  |
| 5–9 | 631 (35.6%) | 0.93 (0.88-0.98) | 0.01 | 0.96 (0.91-1.02) | 0.24 | 0.98 (0.93-1.04) | 0.58 |
| ≥10 | 483 (27.2%) | 1.00 (0.93-1.06) | 0.90 | 1.03 (0.96-1.10) | 0.44 | 1.03 (0.97-1.10) | 0.34 |
| *p for trend* | .. | .. | 0.70 | .. | 0.51 | .. | 0.38 |
| Lower-impact physical activity (hours per week) |  |  |  |  |  |  |  |
| 0 | 1048 (59.2) | 1.00 |  | 1.00 |  | 1.00 |  |
| 1 | 331 (18.7) | 0.95 (0.89-1.01) | 0.11 | 0.97 (0.91-1.03) | 0.29 | 0.97 (0.92-1.04) | 0.42 |
| ≥2 | 390 (22.0) | 0.95 (0.89-1.00) | 0.07 | 0.98 (0.92-1.05) | 0.53 | 0.99 (0.93-1.05) | 0.69 |
| *p for trend* | .. | .. | 0.045 | .. | 0.44 | .. | 0.60 |
| Number of visits to shops and other indoor public places (per week) |  |  |  |  |  |  |  |
| 0 | 237 (13.4) | 1.00 |  | 1.00 |  | 1.00 |  |
| 1-2 | 591 (33.3) | 1.02 (0.94-1.11) | 0.57 | 1.06 (0.98-1.16) | 0.16 | 1.05 (0.97-1.14) | 0.22 |
| 2–4 | 428 (24.1) | 1.08 (0.99-1.18) | 0.07 | 1.12 (1.02-1.22) | 0.02 | 1.12 (1.03-1.22) | 0.01 |
| ≥5 | 518 (29.2) | 1.12 (1.02-1.22) | 0.02 | 1.12 (1.02-1.23) | 0.02 | 1.12 (1.02-1.22) | 0.01 |
| *p for trend* | .. | .. | 0.004 | .. | 0.01 | .. | 0.01 |
| Number of public transport journeys per week |  |  |  |  |  |  |  |
| 0 | 1569 (88.7) | 1.00 |  | 1.00 |  | 1.00 |  |
| 1-5 | 139 (7.9) | 1.05 (0.97-1.15) | 0.24 | 1.04 (0.95-1.13) | 0.44 | 1.02 (0.94-1.11) | 0.60 |
| ≥6 | 60 (3.4) | 1.15 (1.00-1.31) | 0.05 | 1.09 (0.95-1.26) | 0.21 | 1.05 (0.93-1.18) | 0.44 |
| *p for trend* | .. | .. | 0.03 | .. | 0.17 | .. | 0.38 |
| Travel to place of work or study in past week |  |  |  |  |  |  |  |
| No | 697 (39.6%) | 1.00 |  | 1.00 |  | 1.00 |  |
| Yes | 1064 (60.4%) | 1.07 (1.01-1.12) | 0.01 | 1.02 (0.97-1.08) | 0.44 | 1.03 (0.98-1.08) | 0.24 |
| Travel outside of the UK between November 2019, and February 2021<sup>§</sup> |  |  |  |  |  |  |  |
| No | 955 (53.8%) | 1.00 |  | 1.00 |  | 1.00 |  |
| Yes | 602 (33.9%) | 1.08 (1.02-1.13) | 0.004 | 1.11 (1.05-1.16) | <0.001 | 1.09 (1.04-1.15) | 0.001 |
| Multivitamin supplement |  |  |  |  |  |  |  |
| No | 1377 (77.6%) | 1.00 |  | 1.00 |  | 1.00 |  |
| Yes | 397 (22.4%) | 1.08 (1.02-1.16) | 0.01 | 1.08 (1.01-1.15) | 0.02 | 1.09 (1.02-1.16) | 0.009 |
| Iron (only) supplement |  |  |  |  |  |  |  |
| No | 1713 (96.6%) | 1.00 |  | 1.00 |  | 1.00 |  |
| Yes | 61 (3.4%) | 1.17 (0.99-1.38) | 0.07 | 1.16 (0.99-1.37) | 0.07 | 1.15 (0.98-1.35) | 0.09 |
| Zinc (only) supplement |  |  |  |  |  |  |  |
| No | 1673 (94.3%) | 1.00 |  | 1.00 |  | 1.00 |  |
| Yes | 101 (5.7%) | 0.91 (0.83-1.00) | 0.06 | 0.94 (0.85-1.03) | 0.18 | 0.92 (0.84-1.02) | 0.10 |
| Selenium (only) supplement |  |  |  |  |  |  |  |
| No | 1754 (98.9) | 1.00 |  | 1.00 |  | 1.00 |  |
| Yes | 20 (1.1) | 0.87 (0.73-1.03) | 0.10 | 0.86 (0.72-1.03) | 0.11 | 0.86 (0.74-1.01) | 0.07 |
| Fruit, vegetable, or salad intake per day |  |  |  |  |  |  |  |
| 0–2 | 266 (15.1%) | 1.00 |  | 1.00 |  | 1.00 |  |
| 3–4 | 559 (31.7%) | 0.94 (0.87-1.02) | 0.12 | 0.94 (0.86-1.02) | 0.12 | 0.94 (0.87-1.01) | 0.11 |
| 5 | 374 (21.2%) | 0.93 (0.86-1.02) | 0.11 | 0.92 (0.85-1.01) | 0.07 | 0.92 (0.85-1.00) | 0.06 |
| ≥6 | 567 (32.1%) | 0.92 (0.86-1.00) | 0.05 | 0.91 (0.84-0.99) | 0.03 | 0.90 (0.84-0.98) | 0.01 |
| *p for trend* | .. | .. | 0.09 | .. | 0.05 | .. | 0.02 |
| Portions of dairy products or calcium-fortified alternatives per day |  |  |  |  |  |  |  |
| 0–1 | 491 (27.8%) | 1.00 |  | 1.00 |  | 1.00 |  |
| 2 | 485 (27.4%) | 1.07 (1.00-1.14) | 0.04 | 1.08 (1.01-1.15) | 0.02 | 1.09 (1.02-1.16) | 0.01 |
| 3–5 | 407 (23.0%) | 1.10 (1.03-1.19) | 0.01 | 1.12 (1.04-1.20) | 0.002 | 1.10 (1.03-1.18) | 0.01 |
| ≥6 | 386 (21.8%) | 1.06 (0.99-1.14) | 0.08 | 1.09 (1.01-1.17) | 0.03 | 1.08 (1.00-1.16) | 0.04 |
| *p for trend* | .. | .. | 0.04 | .. | 0.01 | .. | 0.02 |
| Self-reported general health |  |  |  |  |  |  |  |
| Excellent | 341 (19.2%) | 1.00 |  | 1.00 |  | 1.00 |  |
| Very good | 701 (39.5%) | 1.03 (0.96-1.10) | 0.42 | 0.99 (0.93-1.06) | 0.83 | 0.99 (0.93-1.06) | 0.80 |
| Good | 474 (26.7%) | 1.02 (0.95-1.09) | 0.67 | 0.98 (0.91-1.06) | 0.61 | 0.97 (0.90-1.05) | 0.46 |
| Fair | 198 (11.2%) | 1.07 (0.97-1.18) | 0.18 | 1.01 (0.91-1.12) | 0.80 | 0.99 (0.90-1.09) | 0.86 |
| Poor | 59 (3.3%) | 1.18 (1.00-1.39) | 0.05 | 1.14 (0.95-1.36) | 0.17 | 1.05 (0.87-1.27) | 0.62 |
| *p for trend* | .. | .. | 0.07 | .. | 0.47 | .. | 0.96 |
| BMI, kg/m<sup>2</sup> |  |  |  |  |  |  |  |
| <25 | 764 (43.2%) | 1.00 |  | 1.00 |  | 1.00 |  |
| 25–30 | 626 (35.4%) | 1.05 (1.00-1.11) | 0.07 | 1.04 (0.98-1.10) | 0.19 | 1.03 (0.98-1.09) | 0.28 |
| >30 | 379 (21.4%) | 1.11 (1.04-1.19) | 0.003 | 1.10 (1.02-1.19) | 0.01 | 1.07 (1.00-1.15) | 0.06 |
| *p for trend* | .. | .. | 0.002 | .. | 0.008 | .. | 0.048 |
| Diabetes or pre-diabetes |  |  |  |  |  |  |  |
| No diabetes | 1636 (92.3%) | 1.00 |  | 1.00 |  | 1.00 |  |
| Pre-diabetes | 52 (2.9%) | 1.03 (0.89-1.19) | 0.70 | 0.97 (0.85-1.11) | 0.65 | 0.99 (0.87-1.13) | 0.88 |
| Type 1 | 12 (0.7%) | 1.00 (0.77-1.30) | 0.99 | 0.95 (0.71-1.27) | 0.73 | 0.99 (0.73-1.34) | 0.96 |
| Type 2 | 72 (4.1%) | 1.14 (0.98-1.32) | 0.09 | 0.97 (0.82-1.15) | 0.71 | 0.96 (0.82-1.12) | 0.59 |
| Asthma/Atopy |  |  |  |  |  |  |  |
| No/No | 1178 (66.4) | 1.00 |  | 1.00 |  | 1.00 |  |
| No/Yes | 305 (17.2) | 0.95 (0.89-1.01) | 0.10 | 0.95 (0.89-1.01) | 0.09 | 0.95 (0.9-1) | 0.07 |
| Yes/No | 145 (8.2) | 1.00 (0.91-1.10) | 0.98 | 0.97 (0.88-1.07) | 0.55 | 0.95 (0.87-1.05) | 0.33 |
| Yes/Yes | 146 (8.2) | 0.99 (0.91-1.08) | 0.82 | 0.99 (0.90-1.09) | 0.87 | 1 (0.91-1.1) | 0.97 |
| COPD |  |  |  |  |  |  |  |
| No | 1728 (97.4%) | 1.00 |  | 1.00 |  | 1.00 |  |
| Yes | 46 (2.6%) | 1.22 (1.01-1.49) | 0.04 | 1.12 (0.93-1.35) | 0.23 | 1.08 (0.91-1.29) | 0.39 |
| Reported feeling anxious or depressed at baseline |  |  |  |  |  |  |  |
| No | 1294 (73.0%) | 1.00 |  | 1.00 |  | 1.00 |  |
| Yes | 479 (27.0%) | 0.95 (0.90-1.00) | 0.07 | 0.92 (0.87-0.97) | 0.003 | 0.91 (0.87-0.97) | 0.002 |
| Anticholinergics |  |  |  |  |  |  |  |
| No | 1685 (95.0%) | 1.00 |  | 1.00 |  | 1.00 |  |
| Yes | 89 (5.0%) | 1.16 (1.01-1.34) | 0.04 | 1.15 (0.99-1.32) | 0.06 | 1.16 (1.00-1.33) | 0.04 |
| Beta blockers |  |  |  |  |  |  |  |
| No | 1659 (93.5%) | 1.00 |  | 1.00 |  | 1.00 |  |
| Yes | 115 (6.5%) | 1.12 (1.01-1.25) | 0.04 | 1.09 (0.98-1.22) | 0.10 | 1.12 (1.01-1.24) | 0.03 |
| Metformin |  |  |  |  |  |  |  |
| No | 1720 (97.0%) | 1.00 |  | 1.00 |  | 1.00 |  |
| Yes | 54 (3.0%) | 1.23 (1.04-1.47) | 0.02 | 1.13 (0.91-1.42) | 0.28 | 1.17 (0.95-1.43) | 0.14 |
| Statins |  |  |  |  |  |  |  |
| No | 1487 (83.8%) | 1.00 |  | 1.00 |  | 1.00 |  |
| Yes | 287 (16.2%) | 1.07 (0.99-1.15) | 0.07 | 1.04 (0.96-1.13) | 0.31 | 1.04 (0.97-1.13) | 0.28 |
| Follow-up duration (days) | .. | 0.001 (0.001-0.001) | <0.001 | 0.001 (0.001-0.002) | <0.001 | 0.001 (0.001-0.001) | <0.001 |
| COVID-19 severity |  |  |  |  |  |  |  |
| Asymptomatic | 1242 (70.6%) | 1.00 |  |  |  | 1.00 |  |
| Symptomatic, not hospitalised | 473 (26.9%) | 1.29 (1.22-1.36) | <0.001 | .. | .. | 1.25 (1.18-1.33) | <0.001 |
| Hospitalised | 44 (2.5%) | 2.13 (1.70-2.67) | <0.000 | .. | .. | 1.92 (1.54-2.40) | <0.000 |
| *p for trend* | .. | .. | <0.001 | .. | .. | .. | <0.001 |

Minimally adjusted models included 1774 participants, except for a few items that included sample sizes were slightly lower (ranged from 1759 to 1773) because of missing values. The fully adjusted analyses included 1707 participants with data available for all factors, and 1694 participants after adjustment for COVID-19 severity. BMI=body-mass index. BTEC=Business and Technology Education Council. COPD=chronic obstructive pulmonary disease. GMR=geometric mean ratio. IMD=index of multiple deprivation.
\*Adjusted for age, sex, and duration of follow-up.
†Adjusted for duration of follow-up and all factors shown except symptom severity.
‡Adjusted for duration of follow-up and all factors shown.
§The 217 participants with unknown or missing travel status were included in the analysis as a separate category to ensure greater power.

**Figure 2:**
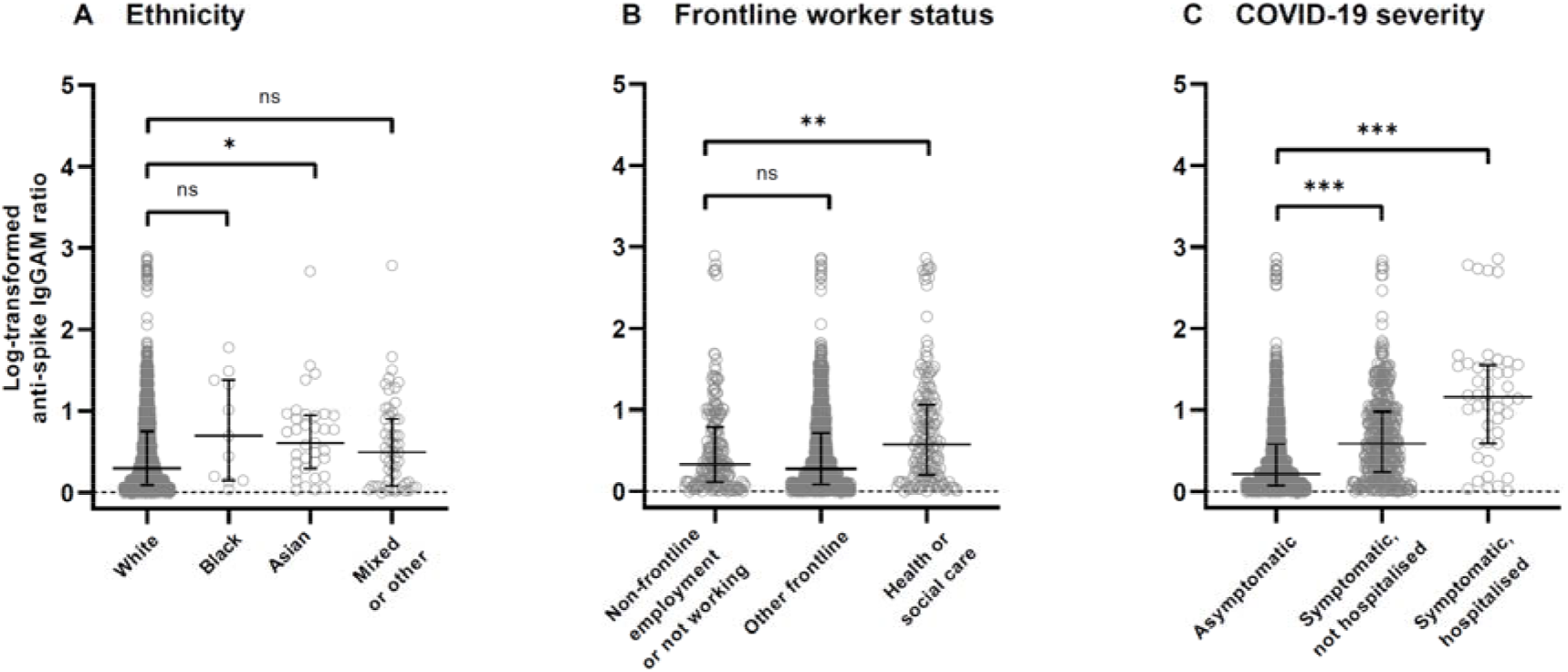
Combined IgG, IgA and IgM anti-S titres in seropositive participants by ethnicity, frontline worker status and COVID-19 severity Log-transformed anti-spike IgGAM ratios are shown for all seropositive participants (n=1774) by ethnic group (A), frontline worker status (B), and COVID-19 severity (C), with horizontal lines showing median and IQR. (A) ‘Black’ indicates people who self-identified their ethnic origin as Black, Black British, African or Caribbean. ‘Asian’ indicates people who self-identified their ethnic origin as Asian or Asian British. ‘Mixed or other’ indicates people who self-identified their ethnic origin as mixed, multiple, or other. (C) COVID-19 severity was classified as ‘asymptomatic’ (non-hospitalised participants who either did not report any symptoms of acute respiratory infection, or whose symptoms were classified as having <50% probability of being due to COVID-19); ‘symptomatic, not hospitalised’ (non-hospitalised participants reporting symptoms of acute respiratory infection that were classified as having ≥50% probability of being due to COVID-19); and ‘hospitalised’ (participants hospitalised for treatment of COVID-19). IgGAM=IgG, IgA, and IgM. *p<0.05. **p<0.01 ***p<0.001. ns = p≥0.05.

The addition of COVID-19 severity to our model of determinants of antibody titres in seropositive participants attenuated the associations observed for BMI and smoking status, but significant associations remained for all other variables (table 3). Inclusion of the severity variable also led to weak associations between antibody titres and male sex (GMR 0.94 [95% CI 0.89–1.00]; p=0.03). Also, use of beta blockers (1.12 [1.01–1.24]; p=0.03) and anticholinergics (1.16 [1.00–1.33]; p=0.04) were positively associated with antibody titres in this model (table 3). p-for-trend analyses suggested lower antibody titres with increasing fruit, vegetable, and salad intake, and higher titres with increasing intake of dairy or calcium-fortified alternatives and increasing age (table 3). The variance in antibody titre explained by the fully adjusted model was increased from 14.5% (R^2^ 0.1447) to 20.5% (R^2^ =0.2047) after including disease severity as a covariate.

## Discussion

In this large, prospective, population-based serological study, we explored determinants of SARS-CoV-2 seropositivity and antibody titres, evaluating more than 80 potential sociodemographic, clinical, and behavioural risk factors. We found that five factors—Asian/Asian British ethnicity, frontline occupation in health or social care, number of visits to shops and other indoor public places, international travel, and high BMI—were strongly associated both with increased risk of SARS-CoV-2 seropositivity among all participants and with higher antibody titres in the subset of seropositive participants. Lower levels of educational attainment and light physical exercise, and higher levels of public transport use and alcohol consumption, were found to associate with increased risk of SARS-CoV-2 seropositivity. Lower intake of dairy or calcium-fortified alternatives and higher intake of fruit, vegetables, and salad were associated with lower antibody titres, as was reporting anxiety or depression. Importantly, most factors associated with antibody titres in seropositive participants retained significance after adjusting for disease severity.

Our results support previous studies that have found increased risk of SARS-CoV-2 seropositivity for healthcare workers,^3,4,12^ people of Asian ethnicity,^3,5,12^ and people with lower educational attainment.^3,4^ Non-White race/ethnicity has previously been highlighted as a determinant of both SARS-CoV-2 seropositivity^23–25^ and antibody titres,^26^ but questions remained over residual confounding.^23^ Despite including a wide range of potential confounders, point estimates for all non-White participants remained elevated in both our seropositivity and titre analyses, and significantly so for Asian/Asian British participants, emphasising the need to further investigate the underlying biological or social factors driving this disparity. While we did not confirm increased seropositivity for Black participants, we lacked statistical power, as they represented only 0.4% of the cohort.

We identified two novel modifiable lifestyle factors associated with SARS-CoV-2 seropositivity: alcohol consumption and light physical exercise. High levels of alcohol intake are known to negatively affect immune response through several mechanisms,^27^ which supports our finding of increased risk among participants consuming more than 15 units of alcohol a week. By contrast, we observed reduced risk among participants doing more than 10 hours of light physical exercise per week. It has been speculated that there is a J-shaped relationship between exercise load and susceptibility to infection, whereby moderate exercise can improve immune response, but prolonged, high-intensity exercise can increase susceptibility to infection.^28^ This curve might explain why we did not see similar benefits for vigorous physical activity.

Our finding that use of vitamin D supplements was associated with increased risk of seropositivity contrasts with a previous study, which found lower risk in a univariable model and no association after adjusting for confounders;^24^ randomised controlled trials are needed to resolve questions around potential effects of vitamin D supplements on susceptibility to SARS-CoV-2. We found no associations for frontline workers not based in health or social care, at odds with previous findings.^11,12^ We also did not observe associations between seropositivity and age or sex, unlike other studies.^5,24,25,29,30^ This may reflect the fact that we adjusted for more potential confounders than most of these studies, and included behaviours that reflect social mixing and influence exposure to infectious index cases.

The strongest association with antibody titre was for disease severity, which explained a further 6% of variatiance when added to our full model, including 35 factors explaining 14.5% of variance in antibody titre together. After adjustment for disease severity, we uncovered ten factors associated with higher titres (greater age, Asian/Asian British ethnicity, higher BMI, working as a frontline worker in a health or care setting, greater number of visits to shops or other indoor places, international travel, taking multivitamin supplements, increased consumption of dairy products or calcium-fortified alternatives, and use of beta blockers and anticholinergic medications) and three associated with lower titres (male sex, high levels of fruit, vegetable, or salad consumption; and reporting feeling anxious or depressed at baseline). Different mechanisms may explain these associations. High intensity and frequency of exposure could be a cause of elevated antibody titres in participants who visited indoor public places or travelled abroad more frequently and in frontline health and social care workers,^31^ supported by previous findings of higher titres in healthcare workers.^14^ Alternatively, greater immune reactivity could be a cause of higher titres, as seen in female participants, who generally have stronger innate and adaptive immune responses than males.^32^ Diet and nutrition are known to affect immune responses^33^ and thus might explain the higher titres observed with use of multivitamin supplements and higher levels of dairy intake (potentially reflecting higher calcium intakes).^34^ However, little evidence is available for the effect of vitamin supplementation in suboptimal rather than micronutrient-deficient diets,^33^ and after adjustment, we found no associations between intake of any individual vitamin supplements and antibody titres. The negative association with fruit, vegetable, and salad consumption was observed for the highest level of intake only (≥6 portions a day); however, despite 40% of vegan or vegetarian participants being included in that category, neither diet type was found to be associated with antibody titres, suggesting it is not the result of a restricted diet. Our finding of a significant positive dose–response relationship between antibody titres and age after adjustment for disease severity supports findings from other studies.^13,35,36^

This study has several strengths. Use of serology to measure SARS-CoV-2 infection reduces collider bias, as serology testing was offered to all participants enrolled in COVIDENCE UK, in contrast to results from external routine testing that had limited availability, particularly at the start of the pandemic. Serology testing has also allowed us to better quantify the risk of SARS-CoV-2 infection by capturing previous infections that were asymptomatic or unconfirmed. A further strength is the use of an assay with high sensitivity and specificity that targets three different types of antibody,^37^ increasing the probability of identifying a past infection. Additionally, we used dried blood spots for our sampling, which have been found to reduce processing failures compared with microtubes,^38^ which are currently used by large seroprevalence surveys.^39^ The prospective nature of our study reduces the potential for reverse causation explaining our findings, and the granularity of our questionnaire allowed us to explore potential determinants and confounders that other studies have not investigated.

Our study also has some limitations. First, COVIDENCE UK is a self-selected cohort, and thus several groups—such as people younger than 30 years, people of lower socioeconomic status, and non-White ethnic groups—are under-represented. This particularly affected our power to investigate outcomes for Black participants, who have been found to be at higher risk of SARS-CoV-2 infection^4,5,12^ and adverse outcomes^23^ than White people. However, insufficient representativeness in a cohort does not preclude identification of causal associations, and self-selection may result in better response to follow-up.^40^ Second, as we included asymptomatic infections in our titre analysis, we were not able to adjust for timing of infection onset, preventing us from capturing the effects of temporal changes in antibody responses. As 40% of our seropositive participants did not experience symptoms suggestive of COVID-19 or provide a symptom onset date, excluding them would have greatly reduced the power and generalisability of our analysis. Third, as with any observational study, we cannot exclude the possibility that some of the associations we report might be explained by residual or unmeasured confounding. For example, the finding that passive smoking but not active smoking was associated with a reduced risk of seropositivity compared with never-smokers should be treated with caution, unless a plausible protective mechanism can be found. However, we have minimised potential confounding by adjusting for a comprehensive list of putative risk factors, and hope that future studies will test for this and other associations demonstrated to determine whether they can be replicated in different populations.

In conclusion, this prospective serological study shows that people of Asian/Asian British ethnicity, frontline workers in health or social care, people with high BMI, and those who had more visits to indoor public places or who had travelled abroad were at higher risk of SARS-CoV-2 seropositivity, after robust adjustment for confounders. Moreover, among seropositive participants, all of these factors associated independently with higher antibody titres, regardless of disease severity. We additionally show that higher alcohol consumption and reduced light physical exercise, both modifiable lifestyle factors, are associated with increased risk of seropositivity. Future research should focus on modifiable risk factors for seropositivity, as well as determinants of antibody titres and other correlates of protection after SARS-CoV-2 infection, to better understand which groups are most at risk of reinfection and what preventive measures might be taken.

## Data Availability

De-identified participant data will be made available upon reasonable request to the corresponding author.

## Contributors

ARM wrote the study protocol, with input from HH, MT, and SOS. HH, MT, JS, SF, GAD, RAL, CJG, FK, AS, and ARM contributed to questionnaire development and design. HH co-ordinated and managed the study, with input from ARM, DAJ, MT, JS, SOS, NP and SM. HH, JS, ARM, and SOS supported recruitment. SF and AGR developed, validated, and performed laboratory assays. MT, HH, MG, and DAJ contributed to data management and coding medication data. MT and GV directly accessed and verified the data. Statistical analyses were done by MT, with input from SOS, ARM, MG, HH, and GV. GV, MT and ARM wrote the first draft of the report. All authors revised it critically for important intellectual content, gave final approval of the version to be published, and agreed to be accountable for all aspects of the work in ensuring that questions related to the accuracy or integrity of any part of the work were appropriately investigated and resolved. MT, HH, DAJ, SOS, ARM, and GV had full access to all data in the study, and ARM had final responsibility for the decision to submit for publication.

## Acknowledgments

This study was supported by a grant from Barts Charity to ARM and CJG (MGU0466). The work was carried out with the support of BREATHE - The Health Data Research Hub for Respiratory Health (MC_PC_19004) in partnership with SAIL Databank. BREATHE is funded through the UK Research and Innovation Industrial Strategy Challenge Fund and delivered through Health Data Research UK.

MT was supported by a grant from the Rosetrees Trust and The Bloom Foundation (M771) until May 2021 and has been supported by the Barts and the London Charity since then. The views expressed are those of the authors and not necessarily those of Barts Charity, BREATHE, or Health Data Research UK. We thank all participants of COVIDENCE UK, and the following organisations who supported study recruitment: Asthma UK, the British Heart Foundation, the British Lung Foundation, the British Obesity Society, Cancer Research UK, Diabetes UK, Future Publishing, Kidney Care UK, Kidney Wales, Mumsnet, the National Kidney Federation, the National Rheumatoid Arthritis Society, the North West London Health Research Register (DISCOVER), Primary Immunodeficiency UK, the Race Equality Foundation, SWM Health, the Terence Higgins Trust, and Vasculitis UK.

Link to the preprint version on medRxiv: https://www.medrxiv.org/content/10.1101/2021.11.02.21265767v1

