## Supplementary Appendix for "Determinants of pre-vaccination antibody responses to SARS-CoV-2: a population-based longitudinal study (COVIDENCE UK)"

### **Supplementary tables**

#### **Table S1. Baseline questions**

| **Sociodemographic** | | | | | | | | | | |
| --- | --- | --- | --- | --- | --- | --- | --- | --- | --- | --- |
| Date of birth (DD/MM/YYYY) |  | | | | | | | | | |
| Post code |  | | | | | | | | | |
| Address |  | | | | | | | | | |
| Please state your **assigned sex at birth.** | • Male  • Female | | | | | | | | | |
| What is your ethnic origin? | • White   - English / Welsh / Scottish / Northern Irish / British - Irish - Gypsy or Irish Traveller - Any other white background   • Mixed / Multiple ethnic groups   - White and Black Caribbean - White and Black African - White and Asian - Any other Mixed / Multiple ethnic backgrounds   • Asian / Asian British   - Indian - Pakistani - Bangladeshi - Chinese - Any other Asian background   • Black / African / Caribbean / Black British   - African - Caribbean - Any other Black / African / Caribbean background   • Arab  • Other Ethnic Group | | | | | | | | | |
| Were you born in the UK? | • Yes  • No | | | | | | | | | |
| Are you a ‘frontline worker’ who has to physically travel to work during lockdown?  Examples include people employed in health and social care, education and childcare, local/national government, food production or sale, the prison service, the police and public transport. | • Yes  • No | | | | | | | | | |
| What is the highest level of education that you have completed? | • Primary school (0)  • Secondary school up to 16 years (1)  • Higher or secondary or further education (A-levels, BTEC, etc.) (2)  • College or university (3)  • Post-graduate degree (4) | | | | | | | | | |
| In the last month, was your household income sufficient to cover the basic needs of your household, such as food and heating? | • Yes  • Mostly  • Sometimes  • No | | | | | | | | | |
| Please select the box that best describes your current housing situation: | • I own my home outright  • I own my home and I am paying a mortgage  • I am renting privately  • I am renting from the council/housing association  • I am staying with friends or family  • I am homeless or living in temporary accommodation  • Other | | | | | | | | | |
| Do you currently claim Universal Credit? | • Yes, I have applied to receive Universal Credit but have **not yet** received any payments  • Yes, I have claimed Universal Credit and received **one or more** payments  • No | | | | | | | | | |
| How many bedrooms are there in your current accommodation? | • 1–10+ | | | | | | | | | |
| Do you live alone? | • Yes  • No | | | | | | | | | |
| How many people other than yourself live in your household? | • Children aged 0–4 years  ▪ 0–10+  • Children aged 5–15 years  ▪ 0–10+  • People aged 16–64 years  ▪ 0–10+  • People aged 65 years or more  ▪ 0–10+ | | | | | | | | | |
| Does your household have any pets? | • Yes  • No | | | | | | | | | |
| Which of the following best describes your current occupational status? | • Employed  • Self-employed  • Retired  • Furloughed  • Unemployed  • Student  • Other | | | | | | | | | |
| Please indicate which types of pet you have at home.  Select all that apply | • Cat  • Dog  • Indoor bird (e.g. budgie, parrot, canary)  • Rabbit / Guinea pig / Hamster  • Tortoise, turtle, lizard or snake  • Other | | | | | | | | | |
| **Behavioural** | | | | | | | | | | |
| In the last week, and on average, how many times per day did you wash your hands with soap and water or use hand sanitiser gel? | • 1–10+ | | | | | | | | | |
| In the last week, how frequently have you worn a face mask while in an indoor public space? | • Always (100% of the time)  • Usually (50–95% of the time)  • Sometimes (1–49% of the time)  • Never (0% of the time)  • Not applicable (I haven’t been in a public space in the last week) | | | | | | | | | |
| In the last week, how frequently have you worn gloves as an infection protection measure while outside the home? | • Always (100% of the time)  • Usually (50–95% of the time)  • Sometimes (1–49% of the time)  • Never (0% of the time)  • Not applicable (I haven’t been in a public space in the last week) | | | | | | | | | |
| Over the last week, how many days did you work/study exclusively from home? | • 0 | • 1 | | • 2 | • 3 | • 4 | | • 5 | • 6 | • 7 |
|  | • NA  • I am not currently working or studying | | | | | | | | | |
| In the last week, how many journeys did you make on public transport? (return trips count as 2 journeys) | • 1–10+ | | | | | | | | | |
| In the last week, how many times have you been inside a shop or supermarket? | • 1–10+ | | | | | | | | | |
| In the last week, how often have you been inside another indoor public space (e.g. café, pub, place of worship, restaurant, gym, day centre, waiting room, school, library, entertainment venue, hairdresser, takeaway restaurant)? | • 1–10+ | | | | | | | | | |
| Did you travel outside of the UK between 1st November 2019 and 31st October 2020? | • Yes  • No | | | | | | | | | |
| Have you travelled outside of the UK since February 2020? | • Yes  • No | | | | | | | | | |
| Over the last week, how many times have you been into the home of someone who does not live in your household?  (We are referring to visits in which you enter inside someone else’s home. Visits where you do not cross the threshold do not count. Neither do visits to the shops or other public places, we will ask about these later.) | • 1–10+ | | | | | | | | | |
| Have you been advised by a doctor or other professional that you should be ‘SHIELDED’ during the coronavirus outbreak?  (‘Shielding’ involves staying at home and minimising face-to-face contact with people outside the home. This advice has been given to people with certain underlying conditions that place them at increased risk of severe illness from COVID-19, including solid organ transplant recipients; people with specific cancers; people with severe respiratory conditions; people with severe combined immunodeficiency (SCID) or homozygous sickle cell disease; people on certain immunosuppressive treatments; and pregnant women with significant heart disease.) | • Yes  • No | | | | | | | | | |
| **Comorbidities** | | | | | | | | | | |
| What is your **current** height? (if you are unsure, please put your best estimate) | • Feet/inches  • Centimetres | | | | | | | | | |
| What is your **current** weight? | • Stones (sts) / pounds (lbs)  • Kilograms (kg) | | | | | | | | | |
| Have you ever been diagnosed with any of the following conditions by a doctor?  Select all that apply. | • Asthma  • Atopic Eczema or Atopic Dermatitis  • Autoimmune disease (e.g. rheumatoid arthritis, multiple sclerosis (MS), lupus (SLE), Crohn’s disease, ulcerative colitis, psoriasis, Raynaud’s disease, scleroderma)  • Cancer  • Cerebral Palsy  • COPD (including chronic bronchitis, and emphysema)  • Cystic Fibrosis  • Dementia  • Diabetes or pre-diabetes  • Hayfever or Allergic Rhinitis  • Heart Attack, Angina or Coronary Artery Disease  • Heart Failure  • High Blood Pressure (Hypertension)  • HIV Infection  • Hyperparathyroidism (overactive parathyroid gland)  • Kidney stones  • Other kidney disease  • Leg Artery Disease (also known as ‘peripheral vascular disease’, ‘peripheral arterial disease’ or ‘intermittent claudication’)  • Mental health disorder  • Motor Neurone Disease  • Organ transplant  • Parkinson's Disease  • Primary immune deficiency (e.g. antibody deficiency, combined immunodeficiency)  • Sarcoidosis  • Sickle Cell Disease (i.e. two copies of altered gene, affected by anaemia and other complications  • Sickle Cell Carrier (also known as ‘sickle cell trait’, with only one copy of altered gene: few symptoms if any)  • Splenectomy (removal of spleen)  • Stroke or Mini-Stroke  • Tuberculosis (TB)  • None of the above | | | | | | | | | |
| You indicated you have been diagnosed with diabetes or pre-diabetes. Please specify your diagnosis: | • Pre-diabetes (high blood sugar levels, not enough to be diagnosed with diabetes)  • Type 1 diabetes  • Type 2 diabetes  • Other type of diabetes | | | | | | | | | |
| Do you currently have cancer? | • Never  • No, cancer cured or in remission  • Yes, currently receiving treatment | | | | | | | | | |
| **[The following three questions on periodontitis were added to the baseline questionnaire from May 20, 2021; participants who were already enrolled were sent the question within the May 20, 2021, monthly follow-up questionnaire]** | | | | | | | | | | |
| Gum disease (periodontal disease, periodontitis) is a common problem with the mouth. People with gum disease might have bleeding gums around the teeth, swollen gums, receding gums, or sore or infected gums with symptoms lasting for at least 2 weeks and not caused by an injury or problems with dentures.  **Do you think you might have gum disease?** | • Yes  • No | | | | | | | | | |
| Have you ever had any adult teeth that became loose on their own, without some injury? (Milk teeth / baby teeth don’t count) | • Yes  • No | | | | | | | | | |
| Have you ever been told by a dental professional that you have lost bone around your teeth? | • Yes  • No | | | | | | | | | |
| Under each heading, please click the ONE box that best describes your health TODAY.  (Anxiety / Depression) | • I am not anxious or depressed  • I am moderately anxious or depressed  • I am extremely anxious or depressed | | | | | | | | | |
| Over the last 12 months, would you say that on the whole, your health has been: | • Excellent  • Very good  • Good  • Fair  • Poor | | | | | | | | | |
| **Vaccination** | | | | | | | | | | |
| Have you ever had the BCG vaccine?    *This is the vaccine against Tuberculosis (TB), it's injected in the upper arm and usually leaves a small scar* | • Yes  • No  • Unsure | | | | | | | | | |
| Have you ever had the MMR vaccine?  *This is the vaccine against measles, mumps and rubella.* | • Yes  • No  • Unsure | | | | | | | | | |
| **Lifestyle** | | | | | | | | | | |
| Which of these best describes your use of cigarettes? | • I have never smoked cigarettes  • I used to smoke cigarettes occasionally but now not at all  • I used to smoke cigarettes daily but now not at all  • I smoke cigarettes occasionally but not every day  • I smoke cigarettes daily | | | | | | | | | |
| Which of these best describes your use of e‑cigarettes (vaping)? | • I have never vaped or used e-cigarettes  • I used to use e-cigarettes occasionally, but now not at all  • I used to use e-cigarettes daily but now not at all  • I vape occasionally but not every day  • I vape daily | | | | | | | | | |
| Are you regularly exposed to smoke from other people’s cigarettes at home or in a car? | • Yes  • No | | | | | | | | | |
| Roughly how many hours have you spent outdoors in the last week? | • 0–10+ hours | | | | | | | | | |
| During the last week, roughly how many hours did you spend doing more vigorous physical exercise of sufficient intensity to make you breathless or to raise your heart rate significantly, such as heavy physical work, more strenuous gardening (e.g. vigorous digging, landscaping) swimming, jogging, aerobics, football, tennis, cycling, gym workout? | • 0–10+ hours | | | | | | | | | |
| During the last week, roughly how many hours did you spend doing lower impact physical exercise to improve flexibility or core strength such as yoga, tai chi or pilates? | • 0–10+ hours | | | | | | | | | |
| During the last week, roughly how many hours did you spend doing light exercise that does not make you particularly breathless, such as light gardening, walking, including walking for pleasure or exercise, walking to the shops, walking to work? | • 0–10+ hours | | | | | | | | | |
| During the past month, how many hours of actual sleep did you get per night on average?  (This may be different than the number of hours you spent in bed) | • 0–24 hours | | | | | | | | | |
| **Diet** | | | | | | | | | | |
| Do you **exclude** any of the following foods from your diet?  Select all that apply. | • Eggs  • Cow’s milk of products made from cow’s milk (e.g. cheese, yoghurt)  • Fish  • White meat (e.g. poultry)  • Red meat  • No, I eat all of these foods | | | | | | | | | |
| Over the **last week**, how many **portions** of the following did you eat **per day**, on average? |  | | | | | | | | | |
| Fruit, vegetables and salad?  *1 portion = 80g (e.g. one apple or two broccoli spears or 3 tablespoons of peas or carrots or one bowl of salad)* | • 0–10+ | | | | | | | | | |
| Dairy products (e.g. cow’s milk, cheese, yoghurts) or calcium-fortified dairy alternatives (e.g. soya milks, soya yoghurts and soya cheeses)  *1 portion = a cup of milk, a standard pot of yogurt or a piece of cheese about the size of two thumbs together (30g).* | • 0–10+ | | | | | | | | | |
| **Oily** fish  e.g. herring, pilchards, salmon, sardines, sprats, trout and mackerel.  *1 portion = a small tin of oily fish (around 100g) or a piece of oily fish about the size of your palm* | • 0–10+ | | | | | | | | | |
| **White fish or seafood**  e.g. cod, haddock, plaice, prawns and tuna  *1 portion = a small (160g) tin of tuna or a piece of white fish about the size of your palm (140g) or 100g prawns* | • 0–10+ | | | | | | | | | |
| Over the last week, how many cups or glasses of fluid did you drink per day, on average?  *1 cup or glass = about 150ml. Non-alcoholic drinks including water, tea, coffee, milk and other soft drinks all count* | • 0–10+ | | | | | | | | | |
| How many units of alcohol did you drink over the last 7 days?  *One unit is ½ a pint (285 ml) of ordinary beer, lager or cider; 25ml of spirits; 1 small glass (75ml) of wine; or 50ml of sherry* | • None  • 1–7 units  • 8–14 units  • 15–21 units  • 22–28 units  • More than 28 units | | | | | | | | | |
| Over the **last month**, have you taken any of the following supplements at least **once per week**?  Select all that apply. | • Multivitamin (including prenatal multivitamins)  • Supplement containing vitamin A only  • Supplement containing vitamin B only  • Supplement containing vitamin C only  • Supplement containing vitamin D only  • Supplement containing calcium only  • Supplement containing calcium and vitamin D combined  • Supplement containing vitamin E only  • Supplement containing zinc only  • Supplement containing iron only  • Supplement containing probiotics  • Supplement containing fish oil, krill oil or other source of omega-3 fatty acids  • Supplement containing cod liver oil  • Supplement containing echinacea  • Supplement containing garlic or garlic powder (allicin)  • Supplement containing turmeric / curcumin  • Supplement containing Cannabidiol (CBD) oil  • Supplements containing folic acid  • Supplement containing Selenium only  • Other (e.g. other micronutrients (such as herbal supplements) or combinations of micronutrients (such vitamin C & zinc)) Please specify:  • None of the above | | | | | | | | | |
| **Medications** | | | | | | | | | | |
| Please type the names of all the medications you are currently taking below, one medication per box. The next pages will collect details about dosage for each.    Include all types of medications taken at home or administered in a hospital or clinic (capsules, tablets, contraceptive pills or implants, inhalers, injections, intravenous infusions, monoclonal antibodies, chemotherapy, immunosuppressants, etc.)  Please note that other pages will collect details about the amount of medicine in each dose (next page) and how often you take each dose (the page after that). If there are any details about your medication that aren’t captured by our form (e.g. if you take different doses of a medicine at different times of day), there will be space to enter them in a blank text box at the end of this section of the questionnaire. |  | | | | | | | | | |
|  |  | |  | | | | **Medication name (generic or brand name, either is fine)** | | | |
|  |  |  | Medication 1 | | | |  | | | |
|  |  |  | Medication 2 | | | |  | | | |
|  |  |  | …. | | | |  | | | |
| Please select the frequency and route that you take your medication: | • Frequency   - 4 times/day - 3 times/day - 2 times/day - One daily - Weekly - Less often than weekly - As needed   • Route   - By mouth - Inhaled - Injected - Other | | | | | | | | | |
| **Recent health** | | | | | | | | | | |
| Since February 1st 2020, have you experienced any of the following symptoms: loss of smell or taste, fever, persistent cough, fatigue, diarrhoea, abdominal pain or loss of appetite? | • Yes, I have had one or more of these symptoms since 1st of February  • No, I have not had any of these symptoms since 1st of February | | | | | | | | | |
| When did your symptoms start?  (DD/MM/YYYY)  *e.g. 25/04/2020* |  | | | | | | | | | |
| Did you have a persistent cough (coughing a lot for more than an hour, or 3 or more coughing episodes in 24 hours)? | • No  • Persistent dry cough (i.e. producing little or no phlegm)  • Persistent productive cough | | | | | | | | | |
| Did you experience unusual fatigue? | • No  • Mild fatigue  • Severe fatigue – I struggled to get out of bed | | | | | | | | | |
| Did you have a loss of sense of smell or taste? | • Yes  • No | | | | | | | | | |
| Did you skip any meals because you felt unwell? | • Yes  • No | | | | | | | | | |
| Since February 1st 2020, have you had a nose/throat swab to test for COVID-19? | • Yes  • No | | | | | | | | | |
| On what date did you have this nose/throat swab?  If you are not sure of the exact date, enter the approximate date (DD/MM/YYYY).  *e.g. 25/04/2020* |  | | | | | | | | | |
| What was the result? |  | | | | | | | | | |

#### **Table S2. Monthly follow-up questions**

| **Questions** | |
| --- | --- |
| Since you last checked in with us, have you had a nose or throat swab for COVID-19 or any other respiratory virus, or has a result from a previous swab test become newly available?  (This question is about tests to detect the virus itself: they are usually done in somebody who has symptoms, but screening of asymptomatic people can also be done. It’s usually a nose/throat swab, but saliva tests are also becoming available) | • Yes  • No |
| On what date did you have this nose / throat swab?  If you are not sure of the exact date, enter the approximate date (DD/MM/YYYY). |  |
| What was the result? Click as many as apply. | • Positive for COVID-19 (SARS-CoV-2 coronavirus)  • Positive for influenza virus  • Positive for another respiratory virus  • Negative for all/any viruses tested  • Not known |
| Since you last checked in with us, have you experienced any of the following symptoms: cold or flu symptoms, sore throat, persistent cough, loss of smell or taste, fever, fatigue, diarrhoea, abdominal pain or loss of appetite? | • Yes, I have had one or more of these symptoms since completing my last COVIDENCE UK questionnaire  • No, I have not had any of these symptoms since completing my last COVIDENCE UK questionnaire |
| When did your symptoms start? (DD/MM/YYYY) |  |
| Did you have a persistent cough (coughing a lot for more than an hour, or 3 or more coughing episodes in 24 hours)? | • No  • Persistent dry cough (i.e. producing little or no phlegm)  • Persistent productive cough |
| Did you experience unusual fatigue? | • No  • Mild fatigue  Severe fatigue – I struggled to get out of bed |
| Did you have a loss of sense of smell or taste? | • Yes  • No |
| Did you skip any meals because you felt unwell? | • Yes  • No |
| Since you last checked in with us, have you travelled outside of the UK? | • Yes  • No |

#### **Table S3. Factors with no evidence of association with seropositivity for COVID-19 in minimally adjusted model**

|  | **Categories** | **Serum positive N (%)** | **OR^*^ (95% CI)** | **P-value** |
| --- | --- | --- | --- | --- |
| **Sociodemographic, occupational, and lifestyle factors** |  |  |  |  |
| Quartiles of IMD rank (reverse) | Q4 | 453 (15.4) | 1.00 |  |
|  | Q3 | 427 (14.9) | 0.94 (0.82-1.09) | 0.43 |
|  | Q2 | 410 (15.2) | 0.95 (0.82-1.10) | 0.50 |
|  | Q1 | 401 (15.5) | 0.93 (0.80-1.08) | 0.32 |
| Household income sufficient for basic needs | Yes | 1585 (15.2) | 1.00 |  |
|  | Mostly | 63 (16.2) | 1.00 (0.76-1.32) | 0.99 |
|  | Sometimes | 10 (12.5) | 0.73 (0.37-1.43) | 0.36 |
|  | No | 38 (15.2) | 0.92 (0.65-1.31) | 0.65 |
| Pregnant | No | 1152 (15.1) | 1.00 |  |
|  | Yes | 1 (11.1) | 0.54 (0.07-4.35) | 0.56 |
| Pre-school children (0-4y) at home with participants | No | 1624 (15.1) | 1.00 |  |
|  | Yes | 69 (19.1) | 1.09 (0.82-1.47) | 0.55 |
| Schoolchildren (5-15y) at home with participants | No | 1468 (14.9) | 1.00 |  |
|  | Yes | 224 (18.3) | 1.04 (0.87-1.25) | 0.66 |
| Cat at home | No | 1323 (15.3) | 1.00 |  |
|  | Yes | 372 (15.2) | 0.96 (0.85-1.09) | 0.57 |
| Dog at home | No | 1293 (15.2) | 1.00 |  |
|  | Yes | 402 (15.3) | 0.96 (0.85-1.08) | 0.50 |
| Feeling anxious or depressed today | No | 1245 (15.2) | 1.00 |  |
|  | Yes | 450 (15.4) | 0.96 (0.85-1.08) | 0.52 |
| Household income sufficient for basic needs | Yes | 1585 (15.2) | 1.00 |  |
|  | No | 111 (15.4) | 0.94 (0.76-1.16) | 0.58 |
| Vigorous physical exercise | 0h | 665 (16.2) | 1.00 |  |
|  | 1-3h | 619 (14.9) | 0.98 (0.87-1.11) | 0.73 |
|  | ≥4h | 409 (14.3) | 0.92 (0.80-1.06) | 0.24 |
| Food choice | None | 1583 (15.1) | 1.00 |  |
|  | Vegetarian | 90 (17.2) | 1.12 (0.89-1.42) | 0.33 |
|  | Vegan | 23 (15.5) | 0.98 (0.62-1.53) | 0.92 |
| Vaping | Never-vaper | 1581 (15.1) | 1.00 |  |
|  | Ex-vaper | 53 (15.6) | 0.95 (0.70-1.28) | 0.73 |
|  | Current vaper | 57 (19.2) | 1.20 (0.89-1.61) | 0.24 |
| BCG vaccinated | No | 208 (15.0) | 1.00 |  |
|  | Yes | 1339 (15.5) | 1.02 (0.87-1.20) | 0.79 |
|  | Unsure | 142 (13.4) | 0.89 (0.71-1.12) | 0.33 |
| Quartiles of fruit, vegetables, and salad intake | Q1 | 248 (16.6) | 1.00 |  |
|  | Q2 | 533 (14.5) | 0.88 (0.74-1.04) | 0.13 |
|  | Q3 | 355 (16.2) | 0.99 (0.83-1.19) | 0.95 |
|  | Q4 | 552 (14.7) | 0.90 (0.77-1.07) | 0.24 |
| Quartiles of total fish intake | Q1 | 297 (16.8) | 1.00 |  |
|  | Q2 | 265 (14.7) | 0.90 (0.75-1.08) | 0.24 |
|  | Q3 | 441 (14.4) | 0.88 (0.75-1.04) | 0.13 |
|  | Q4 | 691 (15.4) | 0.97 (0.83-1.13) | 0.69 |
| Portions of oily fish intake | None | 658 (15.9) | 1.00 |  |
|  | 1/wk | 537 (14.4) | 0.94 (0.83-1.07) | 0.34 |
|  | 2/wk | 334 (15.4) | 1.03 (0.89-1.20) | 0.66 |
|  | 3+/wk | 165 (15.3) | 1.00 (0.83-1.20) | 0.98 |
| Portions of white fish intake | None | 435 (15.7) | 1.00 |  |
|  | 1/wk | 676 (15.0) | 0.99 (0.86-1.13) | 0.83 |
|  | 2/wk | 394 (14.7) | 0.96 (0.83-1.12) | 0.63 |
|  | 3+/wk | 189 (16.3) | 1.08 (0.90-1.31) | 0.41 |
| Cups of non-alcoholic fluids | Q1 | 362 (14.6) | 1.00 |  |
|  | Q2 | 281 (15.1) | 1.04 (0.87-1.23) | 0.69 |
|  | Q3 | 534 (15.3) | 1.05 (0.91-1.21) | 0.53 |
|  | Q4 | 512 (15.7) | 1.02 (0.88-1.19) | 0.75 |
| Actual sleep, h/night | ≤5h | 153 (16.5) | 1.07 (0.89-1.30) | 0.47 |
|  | 6h | 398 (15.0) | 0.96 (0.84-1.10) | 0.59 |
|  | 7h | 702 (15.4) | 1.00 |  |
|  | ≥8h | 442 (14.7) | 0.91 (0.80-1.04) | 0.16 |
| **Supplements** |  |  |  |  |
| Multivitamin supplement | No | 1319 (14.9) | 1.00 |  |
|  | Yes | 377 (16.6) | 1.04 (0.92-1.18) | 0.55 |
| Vitamin A (only) supplement | No | 1688 (15.2) | 1.00 |  |
|  | Yes | 8 (14.3) | 0.85 (0.40-1.80) | 0.67 |
| Vitamin B (only) supplement | No | 1570 (15.1) | 1.00 |  |
|  | Yes | 126 (17.1) | 1.13 (0.93-1.39) | 0.22 |
| Vitamin C (only) supplement | No | 1518 (15.2) | 1.00 |  |
|  | Yes | 178 (16.0) | 1.01 (0.85-1.19) | 0.95 |
| Zinc (only) supplement | No | 1602 (15.1) | 1.00 |  |
|  | Yes | 94 (18.0) | 1.17 (0.93-1.47) | 0.19 |
| Iron (only) supplement | No | 1638 (15.2) | 1.00 |  |
|  | Yes | 58 (16.6) | 1.03 (0.77-1.38) | 0.84 |
| Probiotics supplement | No | 1592 (15.2) | 1.00 |  |
|  | Yes | 104 (15.2) | 0.93 (0.75-1.16) | 0.52 |
| Fish oil, krill oil or other omega-3 supplements | No | 1497 (15.1) | 1.00 |  |
|  | Yes | 199 (16.2) | 1.06 (0.90-1.25) | 0.48 |
| Code liver supplement | No | 1562 (15.2) | 1.00 |  |
|  | Yes | 134 (15.7) | 1.02 (0.84-1.24) | 0.81 |
| Garlic or garlic powder (allicin) supplement | No | 1655 (15.2) | 1.00 |  |
|  | Yes | 41 (18.7) | 1.27 (0.90-1.80) | 0.17 |
| Selenium (only) supplement | No | 1676 (15.2) | 1.00 |  |
|  | Yes | 20 (17.9) | 1.15 (0.71-1.88) | 0.57 |
| **Medical conditions** |  |  |  |  |
| Asthma | No | 1420 (15.2) | 1.00 |  |
|  | Yes | 276 (15.3) | 0.97 (0.85-1.12) | 0.71 |
| Atopy | No | 1256 (15.2) | 1.00 |  |
|  | Yes | 440 (15.4) | 0.97 (0.86-1.09) | 0.61 |
| Asthma/Atopy | No/No | 1121 (15.1) | 1.00 |  |
|  | No/Yes | 299 (15.7) | 1.00 (0.87-1.15) | 0.97 |
|  | Yes/No | 135 (15.9) | 1.04 (0.86-1.27) | 0.68 |
|  | Yes/Yes | 141 (14.8) | 0.92 (0.76-1.11) | 0.38 |
| Heart disease (CAD, HF) | No | 1634 (15.2) | 1.00 |  |
|  | Yes | 62 (15.2) | 1.02 (0.77-1.35) | 0.89 |
| Parkinson’s disease | No | 1693 (15.2) | 1.00 |  |
|  | Yes | 3 (12.5) | 0.81 (0.24-2.73) | 0.73 |
| Arterial disease (IHD, PVD, CVA) | No | 1612 (15.2) | 1.00 |  |
|  | Yes | 84 (15.2) | 1.04 (0.81-1.32) | 0.77 |
| Hypertension | No | 1339 (15.3) | 1.00 |  |
|  | Yes | 357 (15.0) | 1.02 (0.89-1.16) | 0.81 |
| Kidney disease | No | 1667 (15.3) | 1.00 |  |
|  | Yes | 29 (13.6) | 0.86 (0.58-1.29) | 0.47 |
| Major neurological conditions | No | 1655 (15.3) | 1.00 |  |
|  | Yes | 41 (14.3) | 0.94 (0.67-1.32) | 0.73 |
| Cancer | Never | 1556 (15.4) | 1.00 |  |
|  | Past | 131 (13.9) | 0.92 (0.76-1.12) | 0.43 |
|  | Present | 9 (9.9) | 0.61 (0.31-1.23) | 0.17 |
| Immunodeficiency (HIV, PID) | No | 1688 (15.3) | 1.00 |  |
|  | Yes | 8 (12.5) | 0.72 (0.34-1.51) | 0.38 |
| Autoimmune disease (Multiple sclerosis etc.) | No | 1539 (15.1) | 1.00 |  |
|  | Yes | 157 (16.3) | 1.05 (0.87-1.25) | 0.63 |
| Diabetes types | No diabetes | 1567 (15.3) | 1.00 |  |
|  | Pre-diabetes | 49 (14.8) | 1.01 (0.74-1.38) | 0.95 |
|  | Type 1 diabetes | 12 (15.0) | 0.93 (0.50-1.74) | 0.83 |
|  | Type 2 diabetes | 66 (15.2) | 0.99 (0.75-1.30) | 0.93 |
| Periodontitis | No | 1028 (15.2) | 1.00 |  |
|  | Yes | 469 (15.7) | 1.06 (0.94-1.20) | 0.30 |
| General health | Excellent | 333 (14.5) | 1.00 |  |
|  | Very good | 680 (15.3) | 1.08 (0.93-1.24) | 0.31 |
|  | Good | 453 (15.4) | 1.06 (0.91-1.24) | 0.43 |
|  | Fair | 182 (15.9) | 1.06 (0.87-1.29) | 0.55 |
|  | Poor | 47 (14.8) | 0.94 (0.67-1.31) | 0.72 |
| **Medicines** |  |  |  |  |
| Statins | No | 1422 (15.4) | 1.00 |  |
|  | Yes | 274 (14.4) | 0.98 (0.84-1.14) | 0.81 |
| ACE inhibitors | No | 1551 (15.4) | 1.00 |  |
|  | Yes | 145 (13.8) | 0.89 (0.74-1.07) | 0.23 |
| Proton pump inhibitors | No | 1466 (15.2) | 1.00 |  |
|  | Yes | 230 (15.6) | 1.03 (0.89-1.20) | 0.68 |
| Regular inhaled corticosteroids for asthma or COPD | No | 1532 (15.2) | 1.00 |  |
|  | Yes | 164 (16.0) | 1.03 (0.86-1.23) | 0.76 |
| Oral corticosteroids | No | 1648 (15.2) | 1.00 |  |
|  | Yes | 48 (18.4) | 1.22 (0.88-1.67) | 0.23 |
| Systemic Immunosuppressants | No | 1618 (15.2) | 1.00 |  |
|  | Yes | 78 (17.0) | 1.09 (0.85-1.40) | 0.51 |
| SSRIs | No | 1584 (15.3) | 1.00 |  |
|  | Yes | 112 (15.0) | 0.91 (0.74-1.13) | 0.40 |
| non-SSRIs antidepressants | No | 1623 (15.2) | 1.00 |  |
|  | Yes | 73 (15.9) | 1.00 (0.78-1.30) | 0.97 |
| Angiotensin receptor blockers (ARBs) | No | 1595 (15.2) | 1.00 |  |
|  | Yes | 101 (15.7) | 1.07 (0.86-1.34) | 0.56 |
| Vitamin K antagonists | No | 1683 (15.2) | 1.00 |  |
|  | Yes | 13 (20.6) | 1.42 (0.77-2.63) | 0.27 |
| Beta blockers | No | 1586 (15.3) | 1.00 |  |
|  | Yes | 110 (14.4) | 0.94 (0.76-1.16) | 0.56 |
| Thiazides | No | 1649 (15.3) | 1.00 |  |
|  | Yes | 47 (13.9) | 0.94 (0.69-1.29) | 0.71 |
| H2-receptor antagonists | No | 1685 (15.2) | 1.00 |  |
|  | Yes | 11 (14.5) | 0.87 (0.45-1.65) | 0.66 |
| Calcium channel blockers (CCBs) | No | 1532 (15.2) | 1.00 |  |
|  | Yes | 164 (15.3) | 1.05 (0.88-1.26) | 0.60 |
| Beta-2 adrenergic agonists | No | 1528 (15.0) | 1.00 |  |
|  | Yes | 168 (17.6) | 1.15 (0.97-1.38) | 0.11 |
| Anticholinergics | No | 1614 (15.2) | 1.00 |  |
|  | Yes | 82 (16.9) | 1.07 (0.84-1.37) | 0.59 |
| Bronchodilators | No | 1523 (15.0) | 1.00 |  |
|  | Yes | 173 (17.5) | 1.15 (0.96-1.37) | 0.12 |
| NSAIDS | No | 1557 (15.2) | 1.00 |  |
|  | Yes | 139 (15.4) | 1.03 (0.85-1.24) | 0.79 |
| Sodium-glucose co-transporter-2 inhibitors | No | 1687 (15.2) | 1.00 |  |
|  | Yes | 9 (14.8) | 0.87 (0.43-1.78) | 0.71 |
| Anti-platelet drugs | No | 1592 (15.2) | 1.00 |  |
|  | Yes | 104 (15.2) | 1.03 (0.82-1.28) | 0.80 |
| Metformin | No | 1648 (15.2) | 1.00 |  |
|  | Yes | 48 (15.7) | 1.01 (0.73-1.38) | 0.97 |
| Bisphosphonates | No | 1670 (15.2) | 1.00 |  |
|  | Yes | 26 (14.9) | 0.98 (0.64-1.49) | 0.91 |
| Digoxin | No | 1694 (15.2) | 1.00 |  |
|  | Yes | 2 (22.2) | 1.57 (0.32-7.60) | 0.58 |

OR=odds ratio. *Adjusted for age, sex, and duration of follow-up. The sample size ranged from 11,130 to 11,099 for all factors except for periodontitis with 9,768 and pregnancy with 7,655.

#### **Table S4. Seropositivity sensitivity analysis excluding participants with symptom-defined probable COVID-19**

|  | **Categories** | **Serum positive N (%)** | **OR^*^ (95% CI)** | **P-value** |
| --- | --- | --- | --- | --- |
| **Sociodemographic, occupational, and lifestyle factors** |  |  |  |  |
| Age | <30 | 38 (12.6) | 1.00 |  |
|  | 30-39.99 | 82 (14.0) | 1.13 (0.75-1.70) | 0.57 |
|  | 40-49.99 | 190 (17.0) | 1.42 (0.98-2.07) | 0.07 |
|  | 50-59.99 | 321 (13.9) | 1.12 (0.78-1.61) | 0.53 |
|  | 60-69.99 | 465 (12.1) | 0.96 (0.67-1.37) | 0.81 |
|  | 70+ | 281 (13.0) | 1.04 (0.72-1.49) | 0.85 |
| Sex | Female | 937 (13.0) | 1.00 |  |
|  | Male | 440 (14.1) | 1.15 (1.01-1.30) | 0.03 |
| Ethnicity | White | 1310 (13.2) | 1.00 |  |
|  | Mixed/Multiple/Other | 35 (14.1) | 1.06 (0.73-1.52) | 0.76 |
|  | South Asian | 26 (20.3) | 1.61 (1.04-2.50) | 0.03 |
|  | Black/African/ Caribbean/Black British | 6 (15.8) | 1.19 (0.49-2.86) | 0.70 |
| Claiming universal credit | No | 1345 (13.4) | 1.00 |  |
|  | Yes | 26 (10.7) | 0.72 (0.48-1.10) | 0.13 |
| Quartiles of IMD rank (reverse) | Q4 | 358 (13.2) | 1.00 |  |
|  | Q3 | 371 (13.8) | 1.03 (0.88-1.21) | 0.71 |
|  | Q2 | 330 (13.1) | 0.97 (0.83-1.14) | 0.71 |
|  | Q1 | 313 (13.2) | 0.94 (0.80-1.11) | 0.48 |
| Household income sufficient for basic needs | Yes | 1289 (13.3) | 1.00 |  |
|  | Mostly | 51 (15.0) | 1.10 (0.81-1.50) | 0.54 |
|  | Sometimes | 9 (13.0) | 0.95 (0.47-1.92) | 0.88 |
|  | No | 28 (12.7) | 0.91 (0.61-1.36) | 0.65 |
| No. people per bedroom | ≤0.5 | 500 (12.1) | 1.00 |  |
|  | >0.5-0.99 | 394 (13.7) | 1.12 (0.97-1.29) | 0.13 |
|  | 1-1.99 | 442 (14.4) | 1.11 (0.95-1.29) | 0.17 |
|  | 2+ | 29 (13.4) | 1.02 (0.68-1.55) | 0.92 |
| Multi-generational households | Living alone | 217 (11.7) | 1.00 |  |
|  | Single generation | 761 (13.4) | 1.17 (0.99-1.37) | 0.07 |
|  | Two-generation | 383 (14.0) | 1.16 (0.96-1.39) | 0.12 |
|  | Three-generation | 16 (20.0) | 1.81 (1.02-3.20) | 0.04 |
| Highest educational level attained | Primary/Secondary | 164 (14.8) | 1.00 |  |
|  | Higher/further (A levels) | 206 (13.8) | 0.94 (0.75-1.17) | 0.58 |
|  | College | 632 (13.8) | 0.94 (0.78-1.14) | 0.55 |
|  | Post-grad | 374 (11.9) | 0.79 (0.64-0.96) | 0.02 |
| Pregnant | No | 917 (13.0) | 1.00 |  |
|  | Yes | 1 (11.1) | 0.73 (0.09-5.88) | 0.76 |
| Pre-school children (0-4y) at home with participants | No | 1321 (13.2) | 1.00 |  |
|  | Yes | 53 (16.5) | 1.14 (0.82-1.59) | 0.43 |
| Schoolchildren (5-15y) at home with participants | No | 1204 (13.1) | 1.00 |  |
|  | Yes | 169 (15.4) | 1.02 (0.83-1.25) | 0.86 |
| Working-age adult (16-64y) at home with participant? | No | 614 (12.2) | 1.00 |  |
|  | Yes | 759 (14.4) | 1.17 (1.02-1.35) | 0.02 |
| Cat at home | No | 1086 (13.4) | 1.00 |  |
|  | Yes | 290 (12.9) | 0.94 (0.81-1.08) | 0.36 |
| Dog at home | No | 1055 (13.3) | 1.00 |  |
|  | Yes | 321 (13.4) | 0.97 (0.85-1.12) | 0.71 |
| Feeling anxious or depressed today | No | 1013 (13.2) | 1.00 |  |
|  | Yes | 363 (13.8) | 1.01 (0.89-1.16) | 0.84 |
| Household income sufficient for basic needs | Yes | 1289 (13.3) | 1.00 |  |
|  | No | 88 (14.0) | 1.02 (0.80-1.29) | 0.89 |
| Frontline worker | No | 1124 (12.8) | 1.00 |  |
|  | Non-health | 156 (14.3) | 1.13 (0.94-1.37) | 0.19 |
|  | Health | 97 (22.4) | 1.97 (1.55-2.50) | <0.001 |
| Vigorous physical exercise | 0h | 528 (14.1) | 1.00 |  |
|  | 1-3h | 503 (13.0) | 0.97 (0.85-1.11) | 0.71 |
|  | ≥4h | 343 (12.7) | 0.94 (0.81-1.09) | 0.42 |
| Light physical exercise | 0-4h | 487 (14.6) | 1.00 |  |
|  | 5-9h | 509 (14.7) | 1.05 (0.91-1.20) | 0.51 |
|  | ≥10 | 380 (10.8) | 0.77 (0.66-0.89) | 0.001 |
| Lower-impact physical activity | 0 hrs/wk | 801 (13.8) | 1.00 |  |
|  | 1 hrs/wk | 270 (13.5) | 0.98 (0.85-1.14) | 0.81 |
|  | 2+ hrs/wk | 301 (12.1) | 0.87 (0.75-1.00) | 0.06 |
| Food choice | None | 1285 (13.2) | 1.00 |  |
|  | Vegetarian | 73 (15.2) | 1.15 (0.89-1.49) | 0.29 |
|  | Vegan | 19 (13.7) | 0.99 (0.60-1.61) | 0.96 |
| BMI, kg/m2 | <25 | 611 (12.0) | 1.00 |  |
|  | 25-30 | 491 (14.9) | 1.27 (1.11-1.44) | <0.001 |
|  | >30 | 271 (14.1) | 1.16 (0.99-1.35) | 0.07 |
| Smoking | Never-smoker | 756 (12.9) | 1.00 |  |
|  | Ex-smoker | 557 (13.9) | 1.08 (0.96-1.22) | 0.20 |
|  | Current smoker | 64 (13.5) | 0.99 (0.75-1.31) | 0.97 |
| Vaping | Never-vaper | 1282 (13.2) | 1.00 |  |
|  | Ex-vaper | 45 (15.2) | 1.11 (0.80-1.53) | 0.55 |
|  | Current vaper | 45 (16.9) | 1.22 (0.88-1.69) | 0.24 |
| Environmental tobacco smoke (Passive smoking) | No | 1355 (13.4) | 1.00 |  |
|  | Yes | 22 (11.6) | 0.80 (0.51-1.25) | 0.32 |
| BCG vaccinated | No | 169 (13.0) | 1.00 |  |
|  | Yes | 1087 (13.5) | 1.05 (0.88-1.25) | 0.59 |
|  | Unsure | 115 (11.6) | 0.88 (0.68-1.14) | 0.33 |
| No. of public transport journeys /wk | 0 | 1227 (13.3) | 1.00 |  |
|  | 1-5 | 104 (13.2) | 1.14 (0.91-1.41) | 0.26 |
|  | 6+ | 43 (14.9) | 1.27 (0.91-1.77) | 0.16 |
| Travel to work/study in last week | No | 546 (11.6) | 1.00 |  |
|  | Yes | 820 (14.8) | 1.33 (1.17-1.50) | <0.001 |
| Travel outside of the UK Nov2019-Feb2021^†^ | No | 769 (12.6) | 1.00 |  |
|  | Yes | 457 (14.1) | 1.15 (1.02-1.31) | 0.02 |
|  | Unknown/missing | 151 (15.8) | 1.08 (0.89-1.32) | 0.42 |
| Alcohol | None | 342 (12.5) | 1.00 |  |
|  | 1-7 U/wk | 506 (13.8) | 1.15 (0.99-1.33) | 0.06 |
|  | 8-14 U/wk | 256 (12.2) | 1.01 (0.85-1.20) | 0.94 |
|  | 15+ U/wk | 273 (14.9) | 1.24 (1.04-1.48) | 0.02 |
| Quartiles of fruit, vegetables, and salad intake | Q1 | 204 (14.8) | 1.00 |  |
|  | Q2 | 429 (12.6) | 0.86 (0.71-1.03) | 0.10 |
|  | Q3 | 290 (14.3) | 0.99 (0.81-1.20) | 0.90 |
|  | Q4 | 446 (12.7) | 0.87 (0.73-1.05) | 0.14 |
| Portions of dairy products or alternatives intake | 0-1/d | 374 (13.9) | 1.00 |  |
|  | 2/d | 384 (12.6) | 0.90 (0.77-1.05) | 0.19 |
|  | 3-5/d | 314 (12.8) | 0.92 (0.78-1.08) | 0.31 |
|  | 6+/d | 300 (14.2) | 1.04 (0.88-1.23) | 0.63 |
| Quartiles of total fish intake | Q1 | 241 (14.7) | 1.00 |  |
|  | Q2 | 223 (13.3) | 0.92 (0.76-1.12) | 0.41 |
|  | Q3 | 359 (12.6) | 0.87 (0.72-1.04) | 0.11 |
|  | Q4 | 552 (13.3) | 0.94 (0.79-1.11) | 0.47 |
| Portions of oily fish intake | None | 528 (13.7) | 1.00 |  |
|  | 1/wk | 440 (12.7) | 0.95 (0.83-1.09) | 0.47 |
|  | 2/wk | 270 (13.4) | 1.03 (0.87-1.21) | 0.76 |
|  | 3+/wk | 137 (13.6) | 1.01 (0.82-1.24) | 0.93 |
| Portions of white fish intake | None | 357 (13.9) | 1.00 |  |
|  | 1/wk | 557 (13.2) | 0.98 (0.85-1.13) | 0.76 |
|  | 2/wk | 315 (12.7) | 0.93 (0.79-1.10) | 0.38 |
|  | 3+/wk | 146 (13.8) | 1.02 (0.83-1.26) | 0.82 |
| No. of visits to shops and other indoor public place | Q1 | 174 (12.6) | 1.00 |  |
|  | Q2 | 445 (13.3) | 1.14 (0.94-1.38) | 0.18 |
|  | Q3 | 339 (14.1) | 1.33 (1.09-1.63) | 0.01 |
|  | Q4 | 419 (13.1) | 1.37 (1.12-1.68) | 0.003 |
| Cups of non-alcoholic fluids | Q1 | 302 (13.0) | 1.00 |  |
|  | Q2 | 225 (12.9) | 0.99 (0.82-1.19) | 0.90 |
|  | Q3 | 427 (13.1) | 1.01 (0.86-1.18) | 0.90 |
|  | Q4 | 417 (13.9) | 1.03 (0.88-1.21) | 0.68 |
| Actual sleep, h/night | ≤5h | 118 (14.2) | 1.07 (0.86-1.32) | 0.55 |
|  | 6h | 331 (13.5) | 1.00 (0.87-1.16) | 0.96 |
|  | 7h | 571 (13.4) | 1.00 |  |
|  | ≥8h | 356 (12.7) | 0.91 (0.79-1.05) | 0.21 |
| Housing | Owns own home | 834 (12.5) | 1.00 |  |
|  | Mortgage | 371 (15.1) | 1.12 (0.96-1.32) | 0.16 |
|  | Privately Renting | 79 (13.6) | 1.05 (0.80-1.38) | 0.72 |
|  | Renting from council | 44 (15.7) | 1.22 (0.88-1.71) | 0.24 |
|  | Others | 49 (14.0) | 1.10 (0.77-1.56) | 0.60 |
| **Supplements** |  |  |  |  |
| Multivitamin supplement | No | 1075 (13.0) | 1.00 | 0.40 |
|  | Yes | 302 (14.6) | 1.06 (0.92-1.22) |  |
| Vitamin A (only) supplement | No | 1370 (13.3) | 1.00 |  |
|  | Yes | 7 (13.2) | 0.92 (0.41-2.04) | 0.84 |
| Vitamin B (only) supplement | No | 1284 (13.3) | 1.00 |  |
|  | Yes | 93 (14.1) | 1.05 (0.84-1.32) | 0.66 |
| Vitamin C (only) supplement | No | 1239 (13.3) | 1.00 |  |
|  | Yes | 138 (13.8) | 0.98 (0.81-1.19) | 0.87 |
| Vitamin D (alone or with calcium) | No | 873 (12.7) | 1.00 |  |
|  | Yes | 504 (14.6) | 1.14 (1.01-1.29) | 0.03 |
| Zinc (only) supplement | No | 1300 (13.2) | 1.00 |  |
|  | Yes | 77 (16.2) | 1.22 (0.95-1.57) | 0.12 |
| Iron (only) supplement | No | 1327 (13.2) | 1.00 |  |
|  | Yes | 50 (15.7) | 1.15 (0.85-1.57) | 0.36 |
| Probiotics supplement | No | 1292 (13.3) | 1.00 |  |
|  | Yes | 85 (13.8) | 0.99 (0.78-1.25) | 0.92 |
| Fish oil, krill oil or other omega-3 supplements | No | 1222 (13.3) | 1.00 |  |
|  | Yes | 155 (13.6) | 1.01 (0.84-1.21) | 0.92 |
| Code liver supplement | No | 1264 (13.3) | 1.00 |  |
|  | Yes | 113 (14.1) | 1.05 (0.85-1.29) | 0.65 |
| Garlic or garlic powder (allicin) supplement | No | 1344 (13.3) | 1.00 |  |
|  | Yes | 33 (16.6) | 1.27 (0.87-1.85) | 0.22 |
| Selenium (only) supplement | No | 1364 (13.3) | 1.00 |  |
|  | Yes | 13 (13.5) | 0.98 (0.54-1.77) | 0.95 |
| **Medical conditions** |  |  |  |  |
| Asthma | No | 1157 (13.3) | 1.00 |  |
|  | Yes | 220 (13.4) | 0.98 (0.84-1.15) | 0.83 |
| COPD | No | 1345 (13.3) | 1.00 |  |
|  | Yes | 32 (16.6) | 1.31 (0.89-1.93) | 0.17 |
| Asthma/Atopy | No/No | 914 (13.2) | 1.00 |  |
|  | No/Yes | 243 (13.7) | 1.01 (0.87-1.18) | 0.89 |
|  | Yes/No | 109 (13.9) | 1.04 (0.84-1.29) | 0.74 |
|  | Yes/Yes | 111 (13.0) | 0.94 (0.76-1.16) | 0.57 |
| Heart disease (CAD, HF) | No | 1325 (13.3) | 1.00 |  |
|  | Yes | 52 (13.7) | 1.01 (0.75-1.37) | 0.93 |
| Parkinson’s disease | No | 1374 (13.3) | 1.00 |  |
|  | Yes | 3 (12.5) | 0.90 (0.27-3.05) | 0.87 |
| Arterial disease (IHD, PVD, CVA) | No | 1307 (13.3) | 1.00 |  |
|  | Yes | 70 (13.9) | 1.05 (0.81-1.37) | 0.72 |
| Hypertension | No | 1080 (13.3) | 1.00 |  |
|  | Yes | 297 (13.3) | 1.01 (0.88-1.17) | 0.88 |
| Kidney disease | No | 1354 (13.4) | 1.00 |  |
|  | Yes | 23 (11.7) | 0.84 (0.54-1.31) | 0.45 |
| Atopy | No | 1023 (13.3) | 1.00 |  |
|  | Yes | 354 (13.5) | 0.98 (0.86-1.12) | 0.81 |
| Major neurological conditions | No | 1346 (13.3) | 1.00 |  |
|  | Yes | 31 (12.4) | 0.91 (0.62-1.33) | 0.63 |
| Cancer | Never | 1264 (13.5) | 1.00 |  |
|  | Past | 105 (12.0) | 0.90 (0.73-1.12) | 0.34 |
|  | Present | 8 (9.0) | 0.62 (0.30-1.29) | 0.20 |
| Immunodeficiency (HIV, PID) | No | 1372 (13.3) | 1.00 |  |
|  | Yes | 5 (9.3) | 0.60 (0.24-1.52) | 0.28 |
| Autoimmune disease (Multiple sclerosis etc.) | No | 1254 (13.3) | 1.00 |  |
|  | Yes | 123 (14.0) | 1.04 (0.85-1.27) | 0.74 |
| Periodontitis | No | 836 (13.2) | 1.00 |  |
|  | Yes | 381 (13.8) | 1.08 (0.94-1.23) | 0.28 |
| Diabetes types | No diabetes | 1270 (13.3) | 1.00 |  |
|  | Pre-diabetes | 40 (12.9) | 1.00 (0.71-1.40) | 1.00 |
|  | Type 1 diabetes | 12 (15.4) | 1.13 (0.61-2.10) | 0.70 |
|  | Type 2 diabetes | 54 (13.6) | 0.99 (0.73-1.33) | 0.93 |
| General health | Excellent | 279 (12.8) | 1.00 |  |
|  | Very good | 562 (13.4) | 1.07 (0.92-1.25) | 0.40 |
|  | Good | 362 (13.4) | 1.05 (0.89-1.24) | 0.56 |
|  | Fair | 142 (13.8) | 1.06 (0.85-1.31) | 0.63 |
|  | Poor | 31 (12.4) | 0.90 (0.60-1.34) | 0.59 |
| **Medicines** |  |  |  |  |
| Statins | No | 1149 (13.4) | 1.00 |  |
|  | Yes | 228 (12.8) | 0.95 (0.81-1.13) | 0.58 |
| ACE inhibitors | No | 1257 (13.5) | 1.00 |  |
|  | Yes | 120 (12.1) | 0.87 (0.71-1.07) | 0.17 |
| Proton pump inhibitors | No | 1188 (13.2) | 1.00 |  |
|  | Yes | 189 (13.9) | 1.04 (0.88-1.24) | 0.61 |
| Regular inhaled corticosteroids for asthma or COPD | No | 1248 (13.3) | 1.00 |  |
|  | Yes | 129 (14.0) | 1.03 (0.85-1.25) | 0.76 |
| Oral corticosteroids | No | 1339 (13.3) | 1.00 |  |
|  | Yes | 38 (16.1) | 1.21 (0.85-1.72) | 0.30 |
| Systemic Immunosuppressants | No | 1314 (13.3) | 1.00 |  |
|  | Yes | 63 (15.0) | 1.10 (0.83-1.45) | 0.50 |
| SSRIs | No | 1284 (13.3) | 1.00 |  |
|  | Yes | 93 (13.7) | 0.99 (0.79-1.25) | 0.95 |
| non-SSRIs antidepressants | No | 1324 (13.3) | 1.00 |  |
|  | Yes | 53 (13.0) | 0.94 (0.70-1.26) | 0.66 |
| Angiotensin receptor blockers (ARBs) | No | 1296 (13.3) | 1.00 |  |
|  | Yes | 81 (13.7) | 1.04 (0.81-1.33) | 0.75 |
| Vitamin K antagonists | No | 1366 (13.3) | 1.00 |  |
|  | Yes | 11 (18.3) | 1.38 (0.71-2.67) | 0.34 |
| Beta blockers | No | 1287 (13.4) | 1.00 |  |
|  | Yes | 90 (12.6) | 0.92 (0.73-1.16) | 0.48 |
| Thiazides | No | 1343 (13.4) | 1.00 |  |
|  | Yes | 34 (10.7) | 0.79 (0.55-1.14) | 0.21 |
| H2-receptor antagonists | No | 1367 (13.3) | 1.00 |  |
|  | Yes | 10 (15.4) | 1.09 (0.55-2.14) | 0.81 |
| Calcium channel blockers (CCBs) | No | 1240 (13.3) | 1.00 |  |
|  | Yes | 137 (13.6) | 1.04 (0.85-1.26) | 0.72 |
| Beta-2 adrenergic agonists | No | 1243 (13.1) | 1.00 |  |
|  | Yes | 134 (15.6) | 1.17 (0.96-1.42) | 0.11 |
| Anticholinergics | No | 1312 (13.3) | 1.00 |  |
|  | Yes | 65 (14.6) | 1.05 (0.80-1.38) | 0.70 |
| Bronchodilators | No | 1238 (13.1) | 1.00 |  |
|  | Yes | 139 (15.6) | 1.17 (0.97-1.42) | 0.11 |
| NSAIDS | No | 1266 (13.3) | 1.00 |  |
|  | Yes | 111 (13.4) | 1.00 (0.81-1.23) | 0.97 |
| Sodium-glucose co-transporter-2 inhibitors | No | 1372 (13.3) | 1.00 |  |
|  | Yes | 5 (9.8) | 0.62 (0.25-1.57) | 0.32 |
| Anti-platelet drugs | No | 1291 (13.3) | 1.00 |  |
|  | Yes | 86 (13.6) | 1.02 (0.80-1.30) | 0.87 |
| Sex hormone therapy | No | 1267 (13.2) | 1.00 |  |
|  | Yes | 110 (14.6) | 1.13 (0.91-1.41) | 0.25 |
| Paracetamol | No | 1335 (13.4) | 1.00 |  |
|  | Yes | 42 (10.6) | 0.75 (0.54-1.04) | 0.09 |
| Metformin | No | 1337 (13.3) | 1.00 |  |
|  | Yes | 40 (14.2) | 1.02 (0.73-1.44) | 0.89 |
| Bisphosphonates | No | 1356 (13.3) | 1.00 |  |
|  | Yes | 21 (12.6) | 0.93 (0.59-1.49) | 0.77 |
| Digoxin | No | 1375 (13.3) | 1.00 |  |
|  | Yes | 2 (22.2) | 1.74 (0.36-8.45) | 0.49 |
| Vitamin D (OTC or prescribed) | No | 856 (12.6) | 1.00 |  |
|  | Yes | 521 (14.7) | 1.15 (1.02-1.30) | 0.02 |

^*^Adjusted for age, sex, and duration of follow-up.

^†^Participants with unknown or missing travel status were included in the analysis as a separate category to ensure greater power.

BMI=body-mass index. BTEC=Business and Technology Education Council. COPD=chronic obstructive pulmonary disease. OR=odds ratio.

The sample size ranged from 10,334 to 10,229 for all factors except for periodontitis with 9,108 and pregnancy with 7,076. The sensitivity analysis included participants.

#### **Table S5. Factors with no evidence of association with antibody titres in seropositive participants in minimally adjusted model**

|  | **Categories** | **n (%)** | **GMR^*^ (95% CI)** | **P-value** |
| --- | --- | --- | --- | --- |
| **Sociodemographic, occupational, and lifestyle factors** |  |  |  |  |
| Claiming universal credit | No | 1728 (97.8) | 1.00 |  |
|  | Yes | 39 (2.2) | 0.97 (0.83, 1.13) | 0.68 |
| Environmental tobacco smoke (Passive smoking) | No | 1750 (98.7) | 1.00 |  |
|  | Yes | 23 (1.3) | 0.89 (0.76, 1.03) | 0.10 |
| Pregnant | No | 1208 (99.9) | 1.00 |  |
|  | Yes | 1 (0.1) | 1.10 (0.97, 1.25) | 0.13 |
| Pre-school children (0-4y) at home with participants | No | 1696 (95.8) | 1.00 |  |
|  | Yes | 74 (4.2) | 0.96 (0.84, 1.10) | 0.56 |
| Schoolchildren (5-15y) at home with participants | No | 1535 (86.8) | 1.00 |  |
|  | Yes | 234 (13.2) | 1.02 (0.93, 1.11) | 0.70 |
| Working-age adult (16-64y) at home with participant? | No | 743 (42.0) | 1.00 |  |
|  | Yes | 1026 (58.0) | 1.05 (0.99, 1.11) | 0.13 |
| Cat at home | No | 1379 (77.8) | 1.00 |  |
|  | Yes | 394 (22.2) | 1.02 (0.96, 1.08) | 0.60 |
| Dog at home | No | 1347 (76.0) | 1.00 |  |
|  | Yes | 426 (24.0) | 1.03 (0.98, 1.10) | 0.26 |
| Household income sufficient for basic needs | Yes | 1650 (93.0) | 1.00 |  |
|  | No | 124 (7.0) | 0.98 (0.90, 1.06) | 0.58 |
| Food choice | None | 1658 (93.5) | 1.00 |  |
|  | Vegetarian | 92 (5.2) | 0.94 (0.84, 1.06) | 0.33 |
|  | Vegan | 24 (1.4) | 0.88 (0.73, 1.05) | 0.17 |
| Vaping | Never-vaper | 1654 (93.5) | 1.00 |  |
|  | Ex-vaper | 56 (3.2) | 1.12 (0.95, 1.33) | 0.19 |
|  | Current vaper | 59 (3.3) | 1.11 (0.96, 1.29) | 0.17 |
| BCG vaccinated | No | 218 (12.3) | 1.00 |  |
|  | Yes | 1402 (79.3) | 0.99 (0.93, 1.07) | 0.88 |
|  | Unsure | 147 (8.3) | 1.07 (0.95, 1.20) | 0.27 |
| Household income sufficient for basic needs | Yes | 1650 (93.0) | 1.00 |  |
|  | Mostly | 66 (3.7) | 0.94 (0.85, 1.05) | 0.30 |
|  | Sometimes | 15 (0.8) | 1.08 (0.80, 1.45) | 0.63 |
|  | No | 43 (2.4) | 0.99 (0.88, 1.13) | 0.93 |
| Multi-generational households | Living alone | 274 (15.4) | 1.00 |  |
|  | Single generation | 967 (54.5) | 1.00 (0.93, 1.07) | 0.94 |
|  | Two-generation | 515 (29.0) | 1.06 (0.98, 1.15) | 0.13 |
|  | Three-generation | 18 (1.0) | 1.06 (0.85, 1.32) | 0.59 |
| Alcohol | None | 477 (26.9) | 1.00 |  |
|  | 1-7 U/wk | 633 (35.7) | 0.99 (0.93, 1.05) | 0.70 |
|  | 8-14 U/wk | 326 (18.4) | 0.97 (0.91, 1.04) | 0.45 |
|  | 15+ U/wk | 338 (19.1) | 1.02 (0.95, 1.10) | 0.59 |
| Quartiles of total fish intake/wk | Q1 | 312 (17.6) | 1.00 |  |
|  | Q2 | 283 (16.0) | 1.03 (0.95, 1.13) | 0.45 |
|  | Q3 | 458 (25.9) | 1.04 (0.96, 1.12) | 0.33 |
|  | Q4 | 718 (40.5) | 1.05 (0.97, 1.13) | 0.22 |
| Portions of oily fish intake | None | 697 (39.4) | 1.00 |  |
|  | 1/wk | 560 (31.6) | 1.02 (0.97, 1.09) | 0.42 |
|  | 2/wk | 346 (19.5) | 1.03 (0.95, 1.10) | 0.47 |
|  | 3+/wk | 168 (9.5) | 1.04 (0.95, 1.13) | 0.43 |
| Portions of white fish intake | None | 456 (25.7) | 1.00 |  |
|  | 1/wk | 709 (40.0) | 1.03 (0.96, 1.09) | 0.41 |
|  | 2/wk | 410 (23.2) | 1.02 (0.95, 1.09) | 0.61 |
|  | 3+/wk | 196 (11.1) | 1.05 (0.96, 1.14) | 0.29 |
| Cups of non-alcoholic fluids/d | Q1 | 385 (21.8) | 1.00 |  |
|  | Q2 | 293 (16.6) | 0.99 (0.91, 1.07) | 0.80 |
|  | Q3 | 557 (31.5) | 0.97 (0.90, 1.04) | 0.37 |
|  | Q4 | 532 (30.1) | 0.98 (0.91, 1.05) | 0.50 |
| **Supplements** |  |  |  |  |
| Vitamin A (only) supplement | No | 1766 (99.5) | 1.00 |  |
|  | Yes | 8 (0.5) | 1.02 (0.80, 1.30) | 0.89 |
| Vitamin B (only) supplement | No | 1646 (92.8) | 1.00 |  |
|  | Yes | 128 (7.2) | 1.03 (0.93, 1.15) | 0.54 |
| Vitamin C (only) supplement | No | 1588 (89.5) | 1.00 |  |
|  | Yes | 186 (10.5) | 0.97 (0.90, 1.04) | 0.42 |
| Probiotics supplement | No | 1664 (93.8) | 1.00 |  |
|  | Yes | 110 (6.2) | 1.05 (0.94, 1.17) | 0.39 |
| Fish oil, krill oil or other omega-3 supplements | No | 1569 (88.4) | 1.00 |  |
|  | Yes | 205 (11.6) | 1.03 (0.95, 1.13) | 0.47 |
| Code liver supplement | No | 1632 (92.0) | 1.00 |  |
|  | Yes | 142 (8.0) | 1.04 (0.95, 1.14) | 0.40 |
| Garlic or garlic powder (allicin) supplement | No | 1731 (97.6) | 1.00 |  |
|  | Yes | 43 (2.4) | 1.04 (0.87, 1.25) | 0.64 |
| **Medical conditions** |  |  |  |  |
| Asthma | No | 1483 (83.6) | 1.00 |  |
|  | Yes | 291 (16.4) | 1.01 (0.94, 1.08) | 0.86 |
| Atopy | No | 1323 (74.6) | 1.00 |  |
|  | Yes | 451 (25.4) | 0.96 (0.91, 1.01) | 0.16 |
| Heart disease (CAD, HF) | No | 1708 (96.3) | 1.00 |  |
|  | Yes | 66 (3.7) | 1.13 (0.96, 1.32) | 0.13 |
| Arterial disease (IHD, PVD, CVA) | No | 1683 (94.9) | 1.00 |  |
|  | Yes | 91 (5.1) | 1.08 (0.94, 1.23) | 0.27 |
| Hypertension | No | 1401 (79.0) | 1.00 |  |
|  | Yes | 373 (21.0) | 1.00 (0.94, 1.06) | 0.94 |
| Kidney disease | No | 1744 (98.3) | 1.00 |  |
|  | Yes | 30 (1.7) | 0.94 (0.80, 1.10) | 0.44 |
| Major neurological conditions | No | 1730 (97.5) | 1.00 |  |
|  | Yes | 44 (2.5) | 0.95 (0.82, 1.10) | 0.51 |
| Cancer | No | 1633 (92.1) | 1.00 |  |
|  | Yes | 141 (7.9) | 0.99 (0.91, 1.08) | 0.86 |
| Cancer | Never | 1633 (92.1) | 1.00 |  |
|  | Past | 132 (7.4) | 1.00 (0.92, 1.09) | 0.97 |
|  | Present | 9 (0.5) | 0.91 (0.59, 1.39) | 0.66 |
| Immunodeficiency (HIV, PID) | No | 1765 (99.5) | 1.00 |  |
|  | Yes | 9 (0.5) | 1.00 (0.77, 1.30) | 0.99 |
| Autoimmune disease (Multiple sclerosis etc.) | No | 1610 (90.8) | 1.00 |  |
|  | Yes | 164 (9.2) | 0.94 (0.86, 1.02) | 0.11 |
| Periodontitis | No | 1067 (68.6) | 1.00 |  |
|  | Yes | 489 (31.4) | 0.98 (0.93, 1.03) | 0.40 |
| **Medicines** |  |  |  |  |
| ACE inhibitors | No | 1619 (91.3) | 1.00 |  |
|  | Yes | 155 (8.7) | 1.05 (0.97, 1.15) | 0.23 |
| Proton pump inhibitors | No | 1525 (86.0) | 1.00 |  |
|  | Yes | 249 (14.0) | 1.02 (0.94, 1.09) | 0.68 |
| Regular inhaled corticosteroids for asthma or COPD | No | 1596 (90.0) | 1.00 |  |
|  | Yes | 178 (10.0) | 1.04 (0.95, 1.14) | 0.35 |
| Oral corticosteroids | No | 1718 (96.8) | 1.00 |  |
|  | Yes | 56 (3.2) | 0.98 (0.86, 1.11) | 0.72 |
| Systemic Immunosuppressants | No | 1688 (95.2) | 1.00 |  |
|  | Yes | 86 (4.8) | 0.93 (0.84, 1.03) | 0.16 |
| SSRIs | No | 1648 (92.9) | 1.00 |  |
|  | Yes | 126 (7.1) | 1.01 (0.91, 1.12) | 0.86 |
| non-SSRIs antidepressants | No | 1696 (95.6) | 1.00 |  |
|  | Yes | 78 (4.4) | 1.05 (0.94, 1.18) | 0.37 |
| Angiotensin receptor blockers (ARBs) | No | 1669 (94.1) | 1.00 |  |
|  | Yes | 105 (5.9) | 1.04 (0.92, 1.17) | 0.53 |
| Vitamin K antagonists | No | 1761 (99.3) | 1.00 |  |
|  | Yes | 13 (0.7) | 1.01 (0.70, 1.45) | 0.97 |
| Thiazides | No | 1726 (97.3) | 1.00 |  |
|  | Yes | 48 (2.7) | 0.94 (0.83, 1.07) | 0.36 |
| H2-receptor antagonists | No | 1763 (99.4) | 1.00 |  |
|  | Yes | 11 (0.6) | 1.21 (0.94, 1.56) | 0.14 |
| Calcium channel blockers (CCBs) | No | 1602 (90.3) | 1.00 |  |
|  | Yes | 172 (9.7) | 1.00 (0.92, 1.08) | 0.97 |
| Beta-2 adrenergic agonists | No | 1593 (89.8) | 1.00 |  |
|  | Yes | 181 (10.2) | 1.05 (0.96, 1.15) | 0.26 |
| Bronchodilators | No | 1588 (89.5) | 1.00 |  |
|  | Yes | 186 (10.5) | 1.07 (0.97, 1.17) | 0.18 |
| NSAIDS | No | 1626 (91.7) | 1.00 |  |
|  | Yes | 148 (8.3) | 1.05 (0.96, 1.15) | 0.31 |
| Sodium-glucose co-transporter-2 (SGLT2) inhibitors | No | 1765 (99.5) | 1.00 |  |
|  | Yes | 9 (0.5) | 1.51 (0.88, 2.60) | 0.17 |
| Anti-platelet drugs | No | 1666 (93.9) | 1.00 |  |
|  | Yes | 108 (6.1) | 1.08 (0.96, 1.22) | 0.18 |
| Sex hormone therapy | No | 1619 (91.3) | 1.00 |  |
|  | Yes | 155 (8.7) | 1.03 (0.94, 1.13) | 0.52 |
| Paracetamol | No | 1715 (96.7) | 1.00 |  |
|  | Yes | 59 (3.3) | 1.01 (0.86, 1.18) | 0.91 |
| Bisphosphonates | No | 1746 (98.4) | 1.00 |  |
|  | Yes | 28 (1.6) | 1.08 (0.92, 1.27) | 0.36 |
| Digoxin | No | 1772 (99.9) | 1.00 |  |
|  | Yes | 2 (0.1) | 1.16 (0.70, 1.91) | 0.56 |
| Vitamin D (OTC or prescribed) | No | 1111 (62.6) | 1.00 |  |
|  | Yes | 663 (37.4) | 0.96 (0.91, 1.02) | 0.17 |

Minimally adjusted model included 1774 participants except for few items with sample sizes ranged from 1767 to 1773.

BTEC=Business and Technology Education Council. GMR=geometric mean ratio.

^*^Adjusted for age, sex, and duration of follow-up.
